# Using agent-based modeling to compare corrective actions for *Listeria* contamination in produce packinghouses

**DOI:** 10.1101/2022.02.16.22271004

**Authors:** Cecil Barnett-Neefs, Genevieve Sullivan, Claire Zoellner, Martin Wiedmann, Renata Ivanek

## Abstract

The complex environment of a produce packinghouse can facilitate the spread of pathogens such as *Listeria monocytogenes* in unexpected ways. This can lead to finished product contamination and potential foodborne disease cases. There is a need for simulation-based decision support tools that can test different corrective actions and are able to account for a facility’s interior cross-contamination dynamics. Thus, we developed agent-based models of *Listeria* contamination dynamics for two produce packinghouse facilities; agents in the models represented equipment surfaces and employees, and models were parameterized using observations, values from published literature and expert opinion. Once validated with historical data from *Listeria* environmental sampling, each model’s baseline conditions were investigated and used to determine the effectiveness of corrective actions in reducing prevalence of agents contaminated with *Listeria* and concentration of *Listeria* on contaminated agents. Evaluated corrective actions included reducing incoming *Listeria*, modifying cleaning and sanitation strategies, and reducing transmission pathways, and combinations thereof. Analysis of *Listeria* contamination predictions revealed differences between the facilities despite their functional similarities, highlighting that one-size-fits-all approaches may not always be the most effective means for selection of corrective actions in fresh produce packinghouses. Corrective actions targeting *Listeria* introduced in the facility on raw materials, implementing risk-based cleaning and sanitation, and modifying equipment connectivity were shown to be most effective in reducing *Listeria* contamination prevalence. Overall, our results suggest that a well-designed cleaning and sanitation schedule, coupled with good manufacturing practices can be effective in controlling contamination, even if incoming *Listeria* spp. on raw materials cannot be reduced. The presence of water within specific areas was also shown to influence corrective action performance. Our findings support that agent-based models can serve as effective decision support tools in identifying *Listeria-*specific vulnerabilities within individual packinghouses and hence may help reduce risks of food contamination and potential human exposure.

## Introduction

*Listeria monocytogenes* is an environmentally widespread, Gram-positive bacterium known for its ability to grow at refrigeration temperatures (1) and persist in food industry equipment due to the presence of harborage sites and conditions favoring replication of the bacteria (2). Symptoms of *L. monocytogenes* infection can either manifest in the form of relatively lesser signs that include nausea, vomiting, fever and diarrhea, and more severe ones including abortion, meningitis, encephalitis, septicemia and death (1). Though only possessing an incidence rate between 0.1 to 10 cases per 1 million people per year depending on the specific country (3), listeriosis has a case-fatality rate between 20-30% (4), making it a priority in food safety. Within a produce packinghouse facility, introduction of *L. monocytogenes* on incoming raw produce is only one of the contamination routes that needs to be addressed, as re-contamination is possible further along production lines due to the presence of harborage sites within facilities (5). Alternative introduction routes into a facility can include entries via regular staff or equipment movement, or unexpected occurrences (i.e., random events), such as roof leakage due to extreme weather or during specialized equipment repairs. The interplay between product, equipment surfaces, water and employees presents a complicated web of interactions, allowing pathogens like *L. monocytogenes* to spread beyond its initial introduction site to elsewhere within a facility (6). Challenges associated with control of *L. monocytogenes* are further compounded in fresh and ready-to-eat (RTE) foods that do not undergo a kill-step, such as fresh and fresh-cut produce. To combat the risk of contamination, facilities can employ environmental monitoring programs (EMPs) to locate pathogen sources, determine pathogen spread throughout the facility, and verify which control strategies are most effective. EMPs involve the routine collection of sponge and swab samples of strategically selected surfaces within a facility and testing them for *Listeria* spp. as an indicator for conditions that will facilitate *L. monocytogenes* contamination. EMP results play an important role in identifying and implementing control strategies such as cleaning and sanitation programs and hygienic zoning (7), which helps restrict pathogen movement.

A data scarcity due to limited testing as part of a facility’s EMP or low prevalence of *Listeria* spp. positive samples detected as part of the EMP can be supplemented by *in silico* tools for more quantitative analysis. Furthermore, a digital decision support tool can assist when determining which corrective actions to pursue within a facility. Due to the structurally complex nature of these facilities, an agent-based model (ABM) is well-suited to this task thanks to its inherent specialization in modelling the interactions of heterogeneous and autonomous “agents” representing the components of the system under study. Simulating a facility *in silico* can be used for a number of objectives, such as better understanding of pathogen movement, interpreting results of EMPs, and evaluating interventions or capital improvements in the facility. One such tool is “**En**vironmental monitoring with an **A**gent-**B**ased Model of ***L****ist***e***ria*” (EnABLe) (8), which has already been shown to allow for analysis of *Listeria* spp. transmission in the slicing and packaging room of a smoked seafood facility. EnABLe’s flexible systems not only allow for the establishment of a model replica (sometimes referred to as a “digital twin”) of a real food production environment, but the rapid manipulation of any number of model parameters or agent-specific values as well. This inherent modularity can be an incredibly powerful asset in the development and evaluation of different corrective actions. Moreover, the establishment of a model aids in identifying targeted interventions that can be specifically applied to higher risk areas of a facility to mitigate contamination. Thus, the objective of this study was twofold: (i) to construct and validate ABMs for two produce packinghouses using historical sampling data and (ii) use these validated ABMs to quantify the effectiveness of facility-wide and site-specific corrective actions in reducing *Listeria* spp. contamination in wet and dry areas of the packinghouses.

## Methods

Data from two produce packinghouse facilities were used to create an ABM for each facility (models “Facility A” and “Facility B”). ABMs were constructed in NetLogo 6.2.0 (9) following the general structure of the EnABLe model developed by Zoellner et al. (8). The models were run with one-hour time steps for a period of two virtual weeks, with the first week allowing *Listeria* to potentially become introduced and spread in a facility and simulated environmental monitoring (EM) being performed in the second week for model validation. For corrective action scenarios, each corrective action was started from the beginning of the simulation and ran for the entire two weeks. For both facilities, an ABM was constructed of the main room where packing operations were performed. While the two packinghouse facilities have variations in layout and size, both facilities can be broken down into similar production steps as briefly summarized: produce is brought into the facility via crates carried by forklifts and loaded into a flume system. Once loaded, raw produce is then transferred to a cleaning area for culling and waxing. Produce is then sorted according to size and appearance and is directed accordingly to either a reject area or the appropriate hand-packing area (trays and bags in Facility A; only trays in Facility B). Production as well as cleaning and sanitation shifts were modeled based on information provided by the facilities, with Facility A performing weekly cleaning and sanitation separately on two separate days, and Facility B performing daily cleaning and sanitation on each workday as well as extended cleaning and sanitation on Saturday. Both facilities operated on a single shift during workdays (Monday-Friday) with a half hour break in the middle of the shift.

### Model Construction & Specifications

Each model had two types of agents: equipment and employees. Agents’ attributes included the following fixed characteristics:

i. Position (defined by x and y coordinates to represent position in 2D plane)
ii. Distance from floor (i.e., “height” in cm, used to calculate interaction order for agents sharing the same position)
iii. Zone category as per proximity to food products (Zone 1: Food-Contact Surfaces (FCS); Zone 2: Non-Food-Contact Surfaces (NFCS) in close proximity to food and FCS; Zone 3: NFCS not in close proximity to food or FCS (10))
iv. Cleanability, that classified each agent as either (i) cleanable (i.e., any *Listeria* present on an agent could be removed from the agent during a routine cleaning step) or (ii) uncleanable (i.e., an agent that due to its design cannot be effectively cleaned during routine cleaning and thus remains contaminated once *Listeri*a spreads on it)
v. Cleaning frequency
vi. Surface area (cm^2^)

Additionally, each agent had a number of time-varying attributes to track:

i. *Listeria* quantity (both in terms of the absolute number of CFU and concentration per surface area (in CFU/cm^2^) on an agent)
ii. Frequency of contamination from specific sources over the course of the simulation: (a) raw incoming food material, (b) random introduction occurrences that could take place anywhere in the facility, or (c) “Zone 4” (7) introduction (i.e., introduction from areas outside the packing room), which had a more localized effect near actively used doorways
iii. Agent water level (consisting of three levels: 1: no water; 2: damp to the touch; 3: visible water on agent)
iv. Niche formations over the course of the simulation (defined as *Listeria* spreading onto an “uncleanable” agent) or temporary niche formations (defined a contamination of an otherwise “cleanable” agent that remained contaminated after routine cleaning) and how frequently these occur
v. Sampling over the course of the simulation (if the agent has been sampled by the simulated EMP)

Table 1 provides a summary of the modeled agents and their characteristics. Agents were grouped based on their location in the production area (Loading, Cleaning, Sorting, Reject, Bag Packing, Tray Packing, and an Other group that included a collection of agents not fitting elsewhere, such as quality control workstations and computer workstations) and by presence of water within the facility area: “wet” (Loading and Cleaning) and “dry” (Sorting, Bag/Tray Packing, Reject and Other). Following the establishment of the agent list, the contact structure among agents was created by assigning (i) directed and (ii) undirected links. The presence of connections allowed for transfer of *Listeria* from one agent to another, depending on link’s directionality. Directed links representedbone-way connections (termed “contact-links”, consisting of “out-directed-links” on the sending agents and “in-directed-links” on the receiving agents). These directed links provided opportunities for *Listeria* transfer in a single direction (with agents sending *Listeria* performing “transfer”, and agents receiving performing “reception”), such as transfer belts and rollers. Undirected links (termed “proximity-links”) represented repeated contact between two agents, transferring contamination with a certain frequency and regardless of direction. The models were constructed using observations from in-person visits to the modeled facilities by authors C.W.B.-N. and G.S. to conduct behavioral mapping of number and function of workers (11) and to determine layout, key surfaces, water and traffic patterns, and connection pathways (Fig 1). The specifics of produce commodities packed in the two packinghouses and the locations of packinghouses cannot be provided; this was a condition for gaining access into the facilities and their data.

**Table 1.**
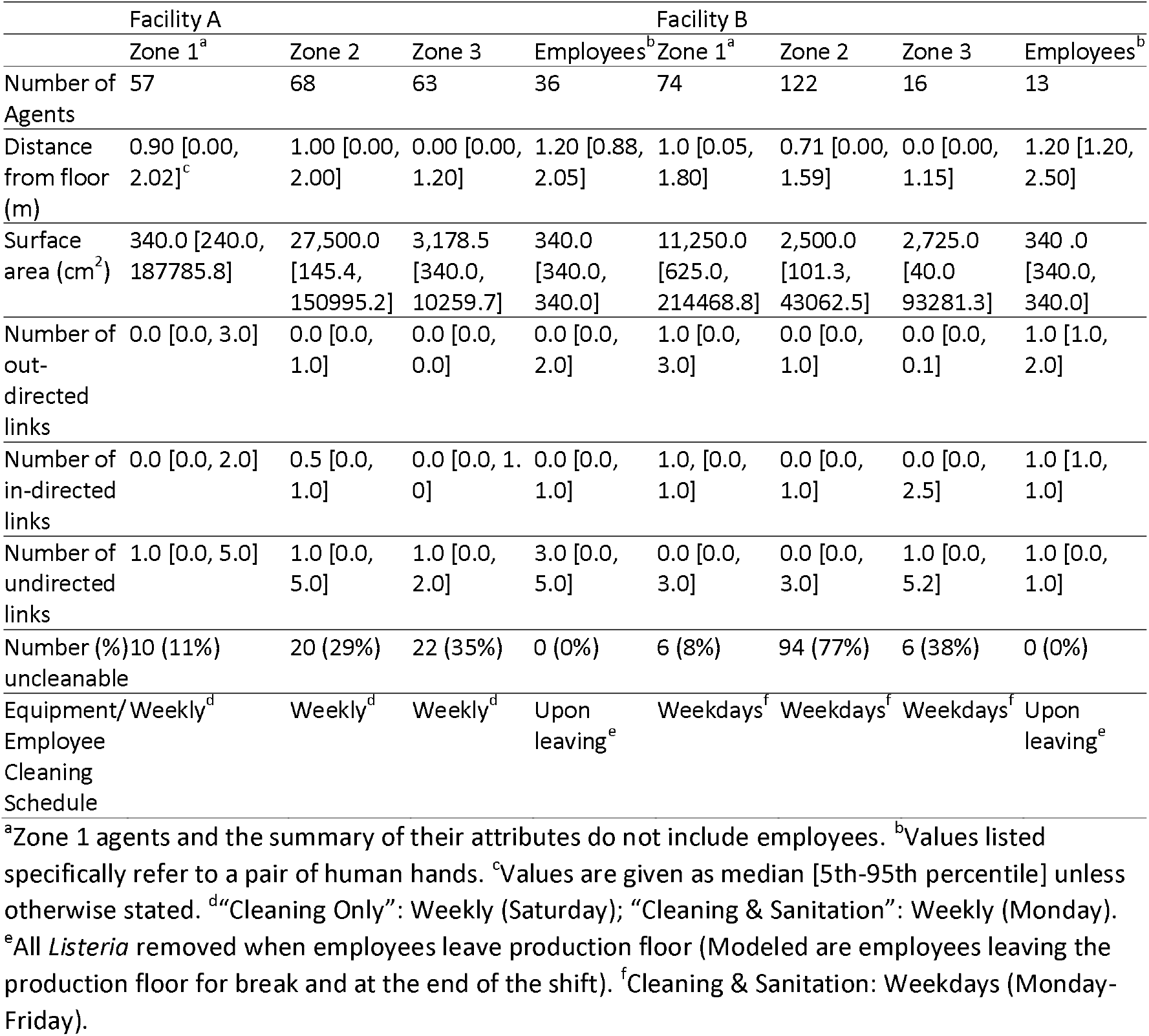
Agent characteristics by zone of agent-based models (Facilities A and B) representing two modeled packinghouses.

**Fig 1.**
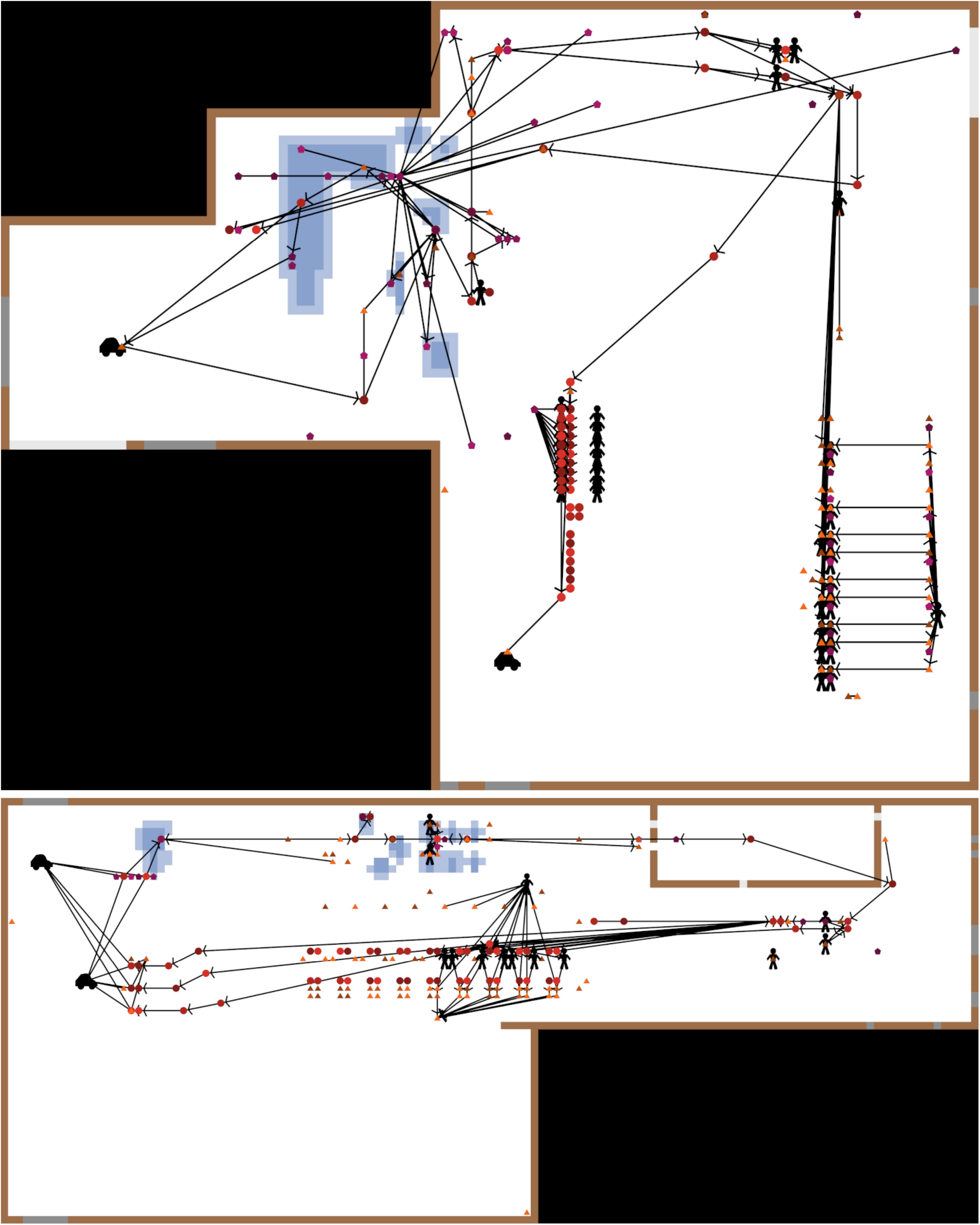
NetLogo views of Facilities A (upper panel) and B (lower panel) illustrating positions of and connections among agents and presence of water at a point in time during production. Circles, triangles and pentagons represent equipment surfaces in zones 1, 2 and 3, respectively; agent water level is denoted by shape color darkness and is independent from floor conditions; employees and forklifts are denoted by specific icons (people and cars respectively); arrows represent the direction of directed agent links and lines without arrows represent undirected links; blue shaded areas represent water presence on the floor (darker colors representing puddles and lighter colors representing damp areas); brown patches denote wall-floor-junctures; grey patches denote doors, with dark grey patches being points of Zone 4 introduction; empty space is denoted by white patches; inactive space not represented by the model is denoted by black.

**Fig 1.**
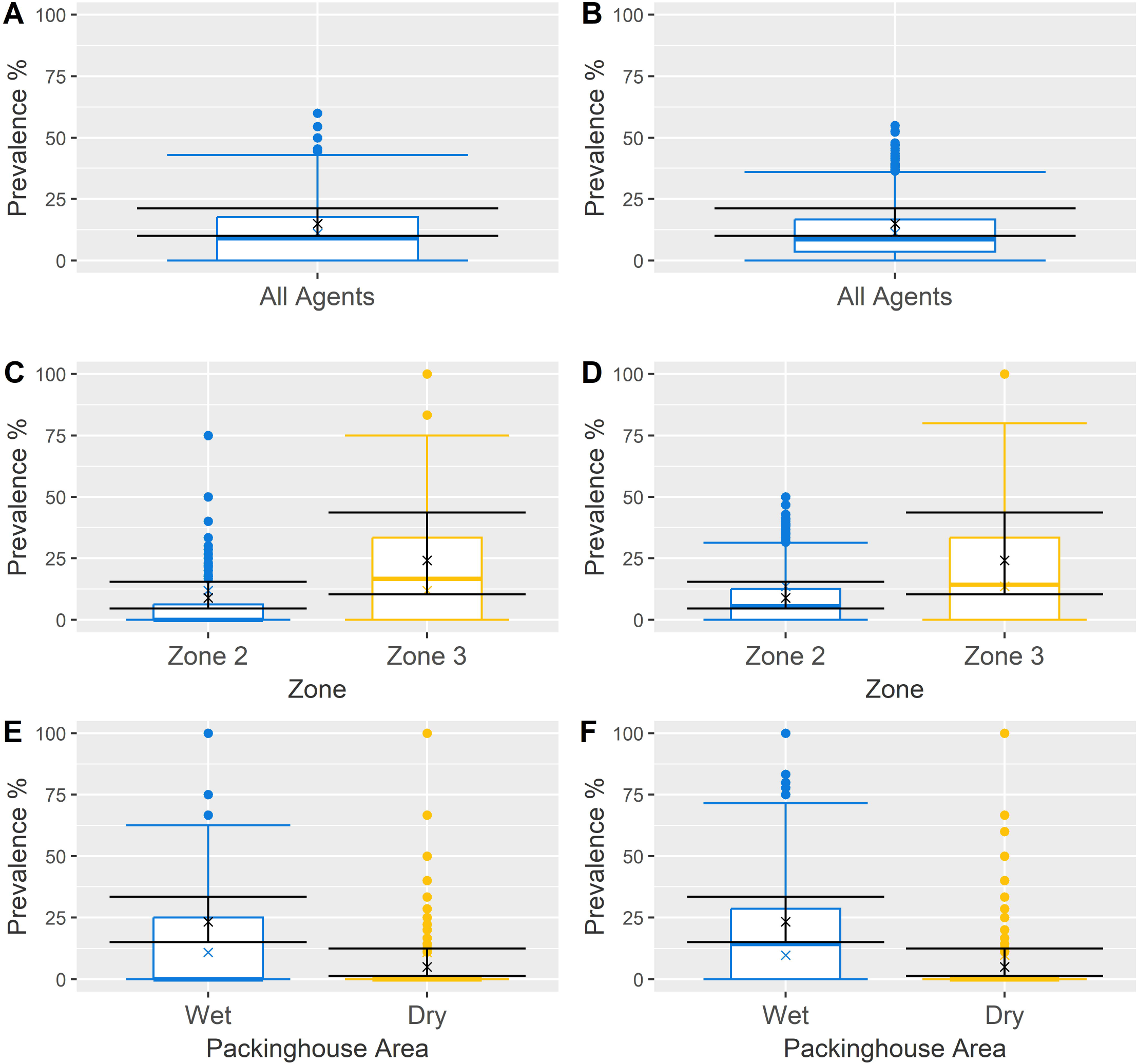
Graphical comparison of baseline Facilities A and B using historical data at midday against simulated sampling results. Results for Facility A and depicted in panels A, C and E, while results for Facility B are in panels B, D and F. Validation groupings investigated included (i) all agents (panels A and B), (ii) Zone category (panels C and D), and (iii) presence of water in the area (panels E and F). Lack of differences between historical (black, covering the mean (denoted with x) and 95% confidence intervals for contamination prevalence) and simulated sampling (colored boxplots, describing the mean (denoted with x), median, interquartile range, 5^th^ and 95^th^ percentiles and any outliers for contamination prevalence) groups indicated the model’s behavior could be considered representative of its respective facility.

A baseline map in the form of text files was first constructed using numerical representation to establish size and grid scale for floor patches, as well as the location of structural components (wall-floor-junctures, open floor, ceiling, and doors). The surface area per patch was 2,500 cm^2^, with Facility A consisting of 109 × 88 patches (9,592) and Facility B consisting of 130 × 56 patches (7,020). Additional maps were then created to represent water level (i.e., none, low, medium, and high) and traffic level (i.e., none, vehicle, low, medium, and high) on the floor for different phases of facility operation. A weekly schedule was also established in the form of a 7×24 csv file for each hour in a week, with each cell detailing the current event for a specific hour (“empty”: no activity; “pre-op”: pre-operations inspection with minimal staff; “production”: standard operations with full staffing and activity; “clean”: “Cleaning Only”/ “Cleaning & Sanitation” operations to remove *Listeria* from equipment) (Tables S1-S5 in S1 File). The schedule was used not only to determine which traffic and water maps to load, but also defined the presence/absence of specific agents (employees and forklifts), as well as *Listeria* introduction processes over time.

Finally, input parameters (as either fixed values or probability distributions) were established from observations and information in the literature to describe *Listeria* growth, transmission, and reduction (Table 2; Tables S6-S9 in S1 File). Data not available in literature sources was acquired from a web-based survey with industry and academic experts that was performed by Sullivan et al. (12) to specifically collect data needed for development of ABMs for fresh produce facilities. Briefly, this expert opinion survey was completed by six individuals (four from academia and two from produce industry backgrounds) with expertise on *Listeria* in food facilities. Each question addressed a specific parameter; survey results were summarized as a median, minimum, and maximum, which were used to parameterize Pert distributions for each parameter alongside a suitable peakedness parameter (Table 2). For each distribution used, its mean and 5^th^-95^th^ percentiles were then calculated to provide summary information. Expert elicitation for the purpose of developing a novel modeling framework is considered beneficial because it permits rapid evaluation of the system and parameter uncertainty, and thus it allows prioritization of future data collection based on the results of sensitivity analysis (13). For model parameters represented as probability distributions, the parameter values across iterations were controlled by a global random seed independent from the rest of the model to ensure a repeatable stream of values was chosen from the distribution between simulations of modeled scenarios. Each iteration within a scenario was also controlled with a local random seed to further ensure repeatability during simulations.

**Table 2.**
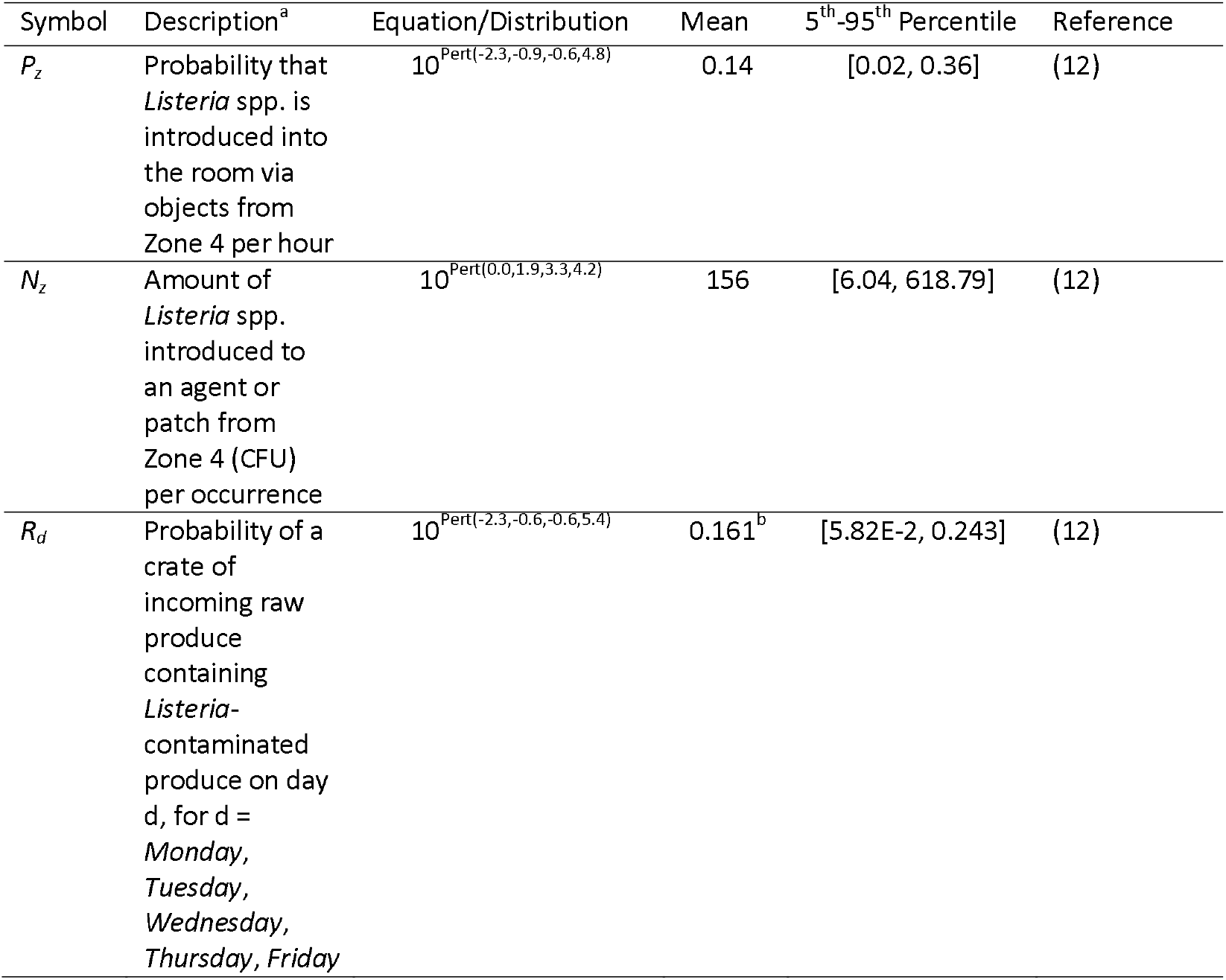
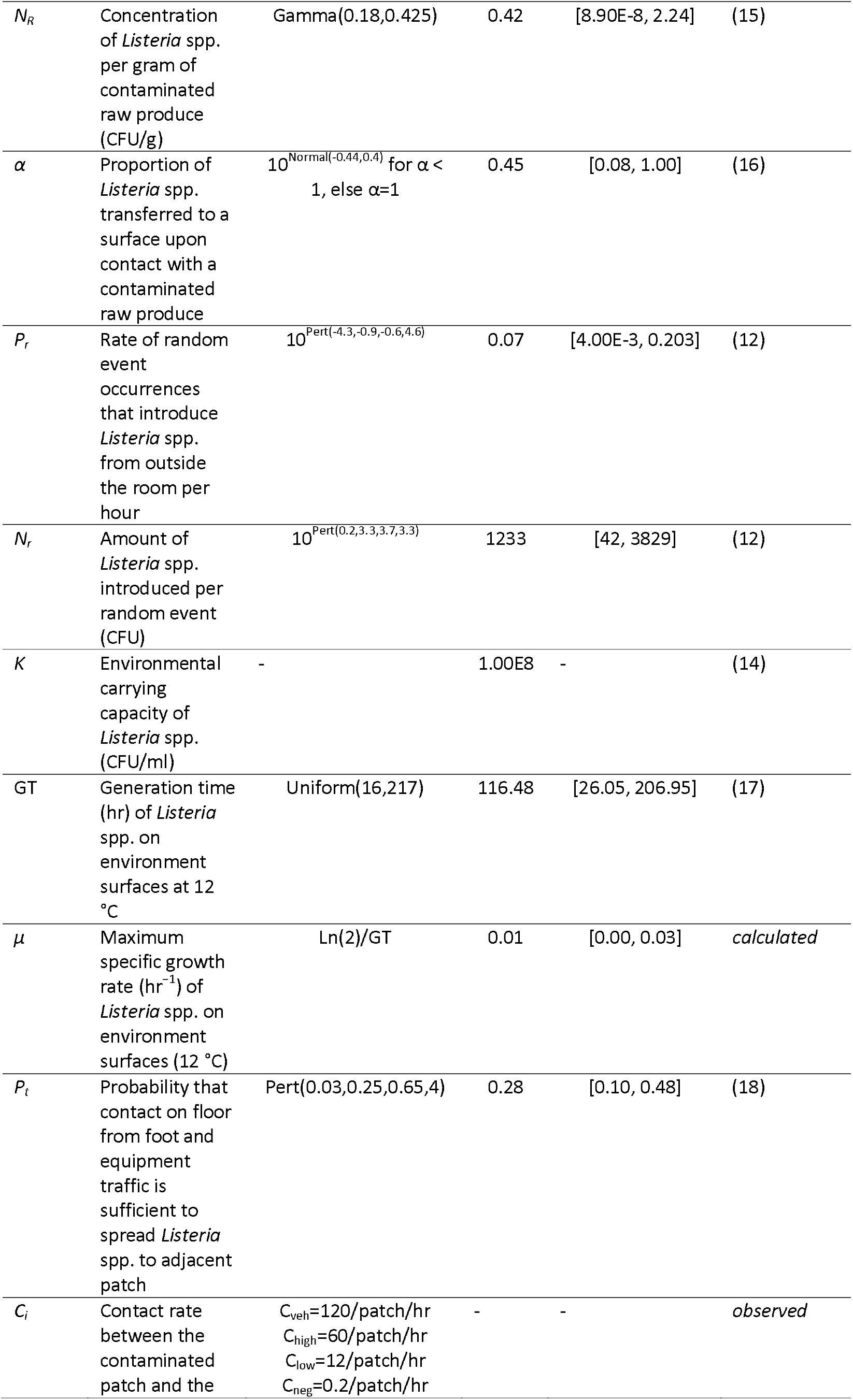
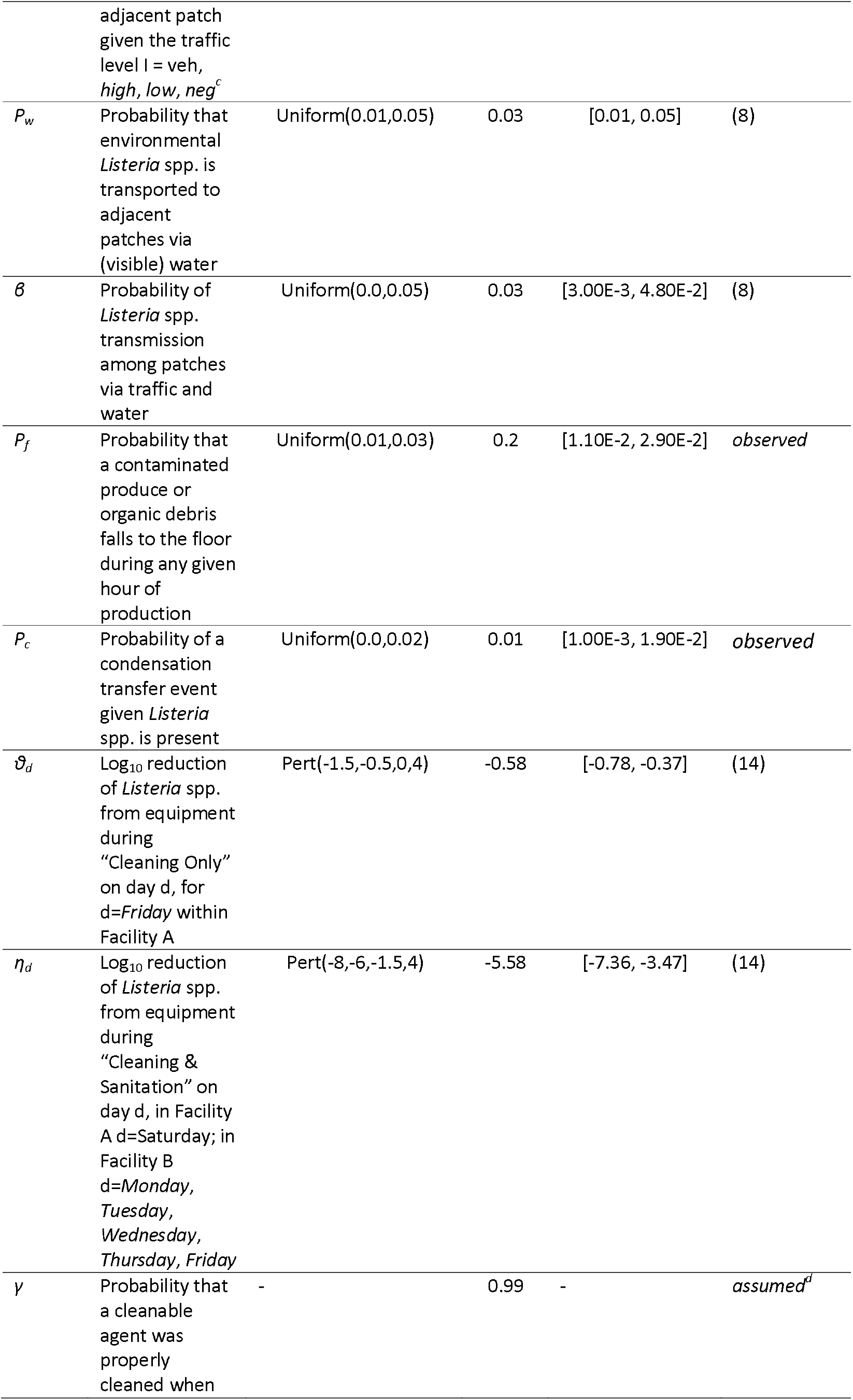
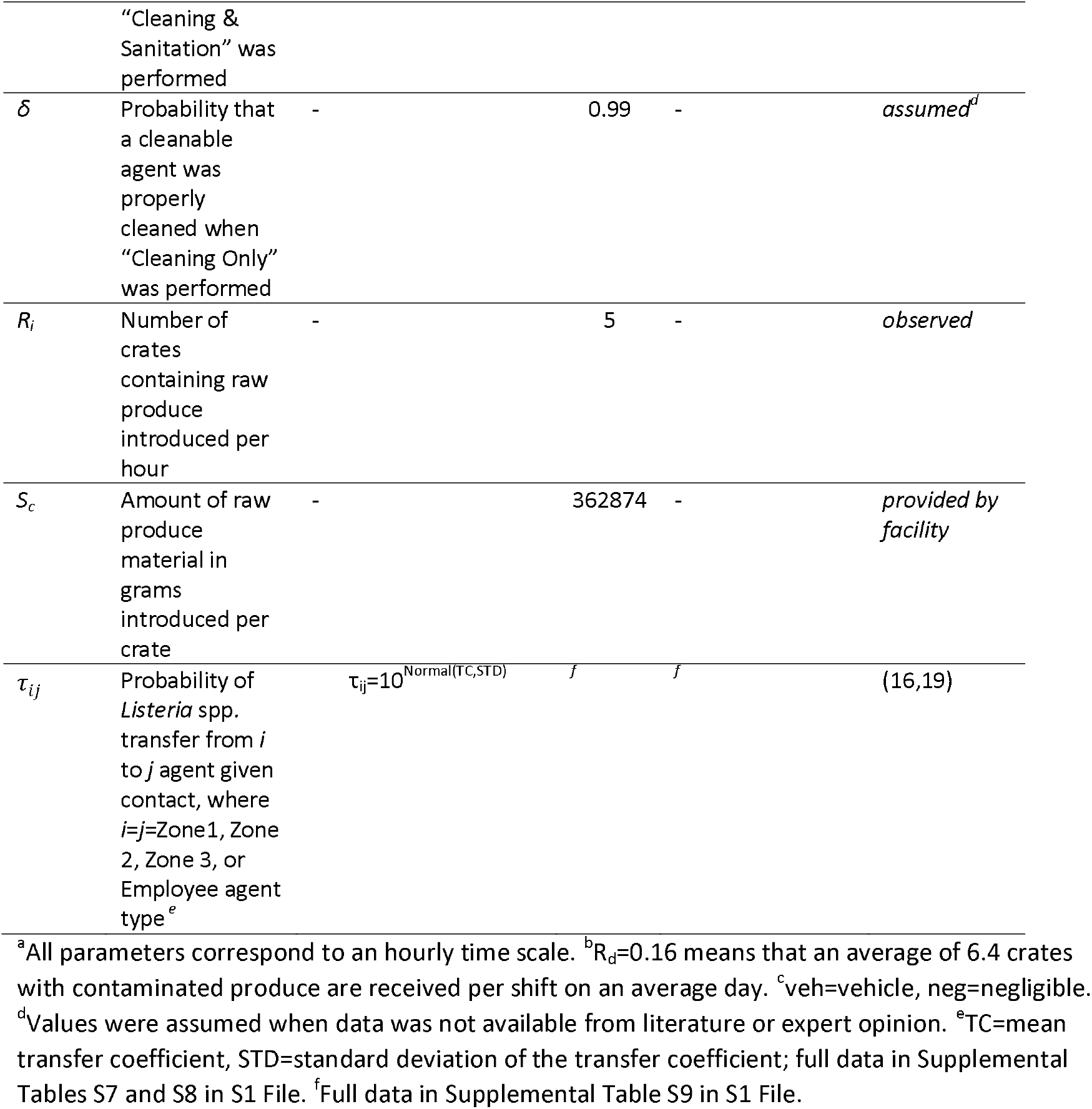
Baseline model input parameters, description, equation and distribution, summary values and sources for *Listeria* spp. introduction, growth, transmission, and reduction.

To determine the degree of reduction in *Listeria* during cleaning for each agent and the timing of these operations, each facility was asked to provide information on the cleaning operations they apply during a regular week. These cleaning activities were modeled as two different levels of reduction: “Cleaning Only” (average of 0.5 log_10_ *Listeria* reduction) and “Cleaning & Sanitation” (average of 6 log_10_ *Listeria* reduction) (14) (for details see S1 File).

Several assumptions were made in the model for simplicity: firstly, temperature was uniform and constant at 12°C within the facility and external weather conditions were not accounted for. Secondly, *Listeria* on the floor (i.e., patches) was not picked up by agents (i.e., modeled equipment surfaces and employee hands) due to a lack of data regarding frequency of occurrence and amount of *Listeria* transferred, as well as a lack of mechanics within the model to adequately judge if an agent’s surface height is too far from the floor to become directly contaminated from the floor. Additionally, in the model employees were allocated to their working stations, however, their movement around the facility was represented through the traffic map and it was assumed they did not deviate from these patterns. This assumption was not expected to have affected the model prediction. Employees driving forklifts were not accounted for in the model because they do not contact the floor or agents in the model system. The value of the probability of a cleanable agent being properly cleaned either during “Cleaning & Sanitation” (γ) or “Cleaning Only” (δ) was assumed due to lack of information. To determine the potential impact of the assumption we tested smaller values of these probabilities but the impact was negligible (Figure S6) and thus the assumed value was considered acceptable. Finally, an agent’s cleanability being switched from “uncleanable” to “cleanable” in scenarios simulating corrective actions was assumed to either represent (i) the inclusion of equipment that was not previously cleaned on a regular basis into facility’s regular cleaning and sanitation operations schedule or (ii) modification or replacement of previously difficult to clean equipment to allow it to be fully cleaned during regular cleaning operations; thereby, in the model a previously uncleanable agent representing such equipment became cleanable.

The models described above, and defined with parameters in Table 2 (and Tables S6-S9 in S1 File), were considered as the “baseline model” for Facilities A and B. All statistical analyses of data generated through model simulations were conducted in R 4.0.5 (20) using the ‘data.table’ package (21) to import large files. To aid interpretation and comparison of results from different sensitivity and scenario analyses, in each model iteration two primary outcomes of interest were recorded at Midday (12:00 pm) on Wednesday of the second simulation week (which in the model was coded as the last action before the mid-shift break). This timing for reporting of the model outcomes was selected to allow for the observation of employee contamination levels just prior to going on break while equipment-related data was collected, mimicking the sampling methods in the historical EM data used for validation. The two outcomes of interest were: (i) the prevalence of contaminated agents (*P_W*) and (ii) *Listeria* concentration on contaminated agents (*C_W*, CFU/cm^2^). The outcome *P_W* was calculated by first estimating the prevalence of contaminated agents in one iteration (overall or in a specific subset of agents) and then to summarize prevalence across model iterations we used a boxplot; the median was recorded for comparisons. Similarly, the outcome *C_W* was summarized over agents (overall or in a specific subset of agents) and iterations using a boxplot; the median was recorded for comparisons.

### Validation and Verification

Both models were validated using historical EM data collected from the respective facility and by recreating analogous *in silico* sampling scenarios that targeted the same equipment surfaces within the model. Historical EM data regarding *Listeria* presence throughout each facility was collected from a complementary study (22), in which Zone 2 and 3 surfaces were sampled using individually packaged sponges hydrated with 10 mL Dey-Engley neutralizing buffer. On a given day, sampling was performed by collecting 3-36 samples in Facility A, and 19-30 samples in Facility B; samples were collected 3-4 hours into a facility’s production cycle and tested for the presence of *Listeria* spp. using the Food and Drug Administration Bacteriological Analytical Manual method (23). A simulated sampling routine was performed in each model using the sampling schedule and the number of samples used for collection of the historical data. Simulated sampling was weighted to favor sites that were historically more often sampled; the weight was calculated by dividing the individual agent’s sampling probability by the sum of sampling probabilities of all agents in historical data:

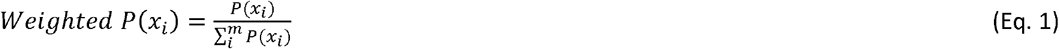

where *x*_i_ is an individual agent among a total of *m* agents and *P* stands for probability.

Simulated environmental sampling was interpreted with an assumed false negative rate of 10% if the agent’s *Listeria* concentration was ≤10 CFU/cm^2^, and 1% if the contamination level was between 11-100 CFU/cm^2^. Samples from agents with a *Listeria* concentration over 100 CFU/cm^2^ were assumed to have a zero false negative rate (8). Each model (Facility A and B) was used to run a 1,000-iteration BehaviorSpace experiment in NetLogo to determine the contamination status of agents representing historical sampling data and then compare model predictions with historical sampling data for the same sampled surfaces. Validation data were evaluated graphically and using a Chi Square Test, or Fisher’s Exact Test (if the number of samples in the group were too small for use of Chi Square test), to determine if prevalence of positive samples in historical data was statistically significantly different from the mean prevalence of positive samples predicted by the model; these comparisons were conducted by shift, zone classification and wet/dry area type. The 95% confidence intervals (95% CI) for the historical prevalences were estimated using the Wilson score interval method in the ‘Hmisc’ R package (24) to describe the level of certainty in the mean estimates while accounting for the sampling effort. Next, we graphically determined whether the 95% CI for the historical data fall within the range of prevalences predicted by the model (described by boxplots).

Each model was additionally verified to be functioning correctly using NetLogo’s own debugging tool for code integrity. Model mechanics were tested using extreme scenarios and simplified models that only ran isolated parts of the original systems. Models for both facilities were also tested on alternate hardware to ensure they remained functional on other computer systems.

### Sensitivity Analysis

A 2-sided partial rank correlation coefficient (PRCC) evaluation using the ‘epiR’ R package (25) was used to perform a sensitivity analysis to identify relationships between the predicted agent contamination prevalence at Midday Week 2 of a randomly selected day from among the days when the facility underwent EM in historical data (Facility A: Monday, Tuesday, and Thursday; Facility B: Monday, Tuesday, and Wednesday) and specific input parameters. Coefficients were then filtered against a Bonferroni-corrected significance level (p=0.05/46=0.0011 and p=0.05/45=0.0011 for Facility A and B, respectively).

### Scenario Analysis

Scenario analyses evaluated the effect of several corrective actions that were created to simulate targeted control and cleaning strategies (Table 3). In Facility B, regular “Cleaning & Sanitation” of equipment is performed daily and thus these corrective actions were already embedded in the baseline model configuration of Facility B. However, Facility A performs both “Cleaning Only” and “Cleaning & Sanitation” procedures only once a week (Saturdays and Mondays, respectively), therefore scenario analysis for Facility A’s risk-based corrective actions additionally tested both (i) daily “Cleaning Only” of equipment and (ii) daily “Cleaning & Sanitation”. In scenarios involving modification of the Master Sanitization Schedule, agents previously designated as “uncleanable” (if that was because they were never cleaned according to the schedule of cleaning in the baseline model) could now be eligible to undergo cleaning (thus, their cleanability attribute would be changed to “cleanable”), depending on their respective predicted mean contamination probability in the baseline model. Scenario AI_04 (“Transmission Pathways Modification Corrective Action”) involved the modification of connections between specific agents to represent corrective actions that had minimal impact to facility function and a higher likelihood of implementation by a facility in response to a contamination event. This criterion was determined by (i) identifying changes in the model that would involve the smallest number of modifications of agent-to-agent connections and (ii) considering the practical feasibility of making those changes in a packinghouse.

**Table 3.**
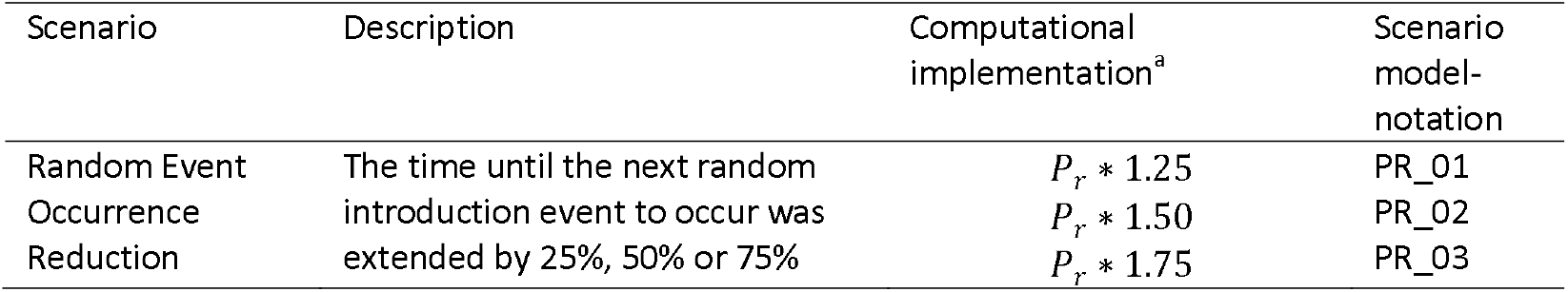
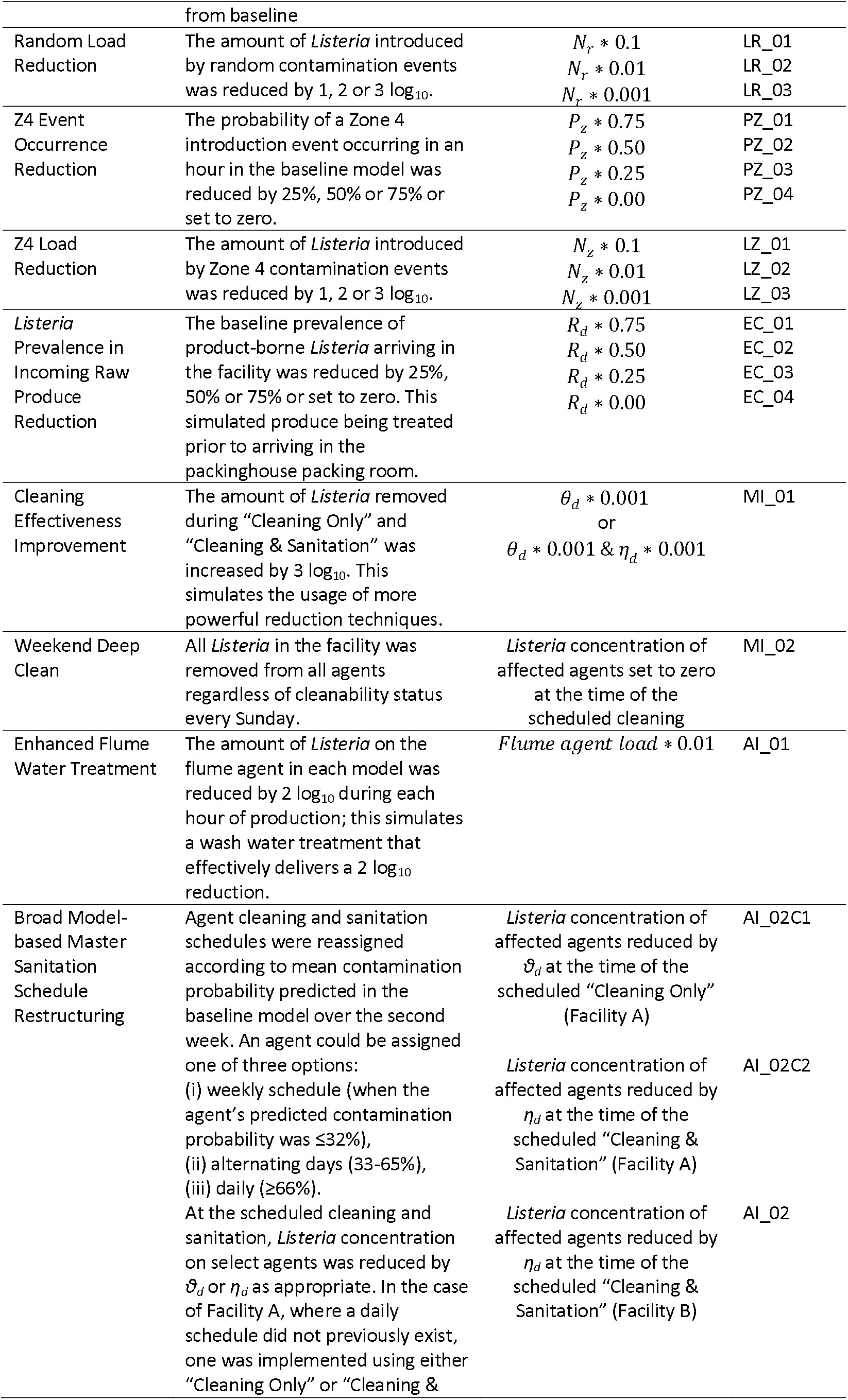
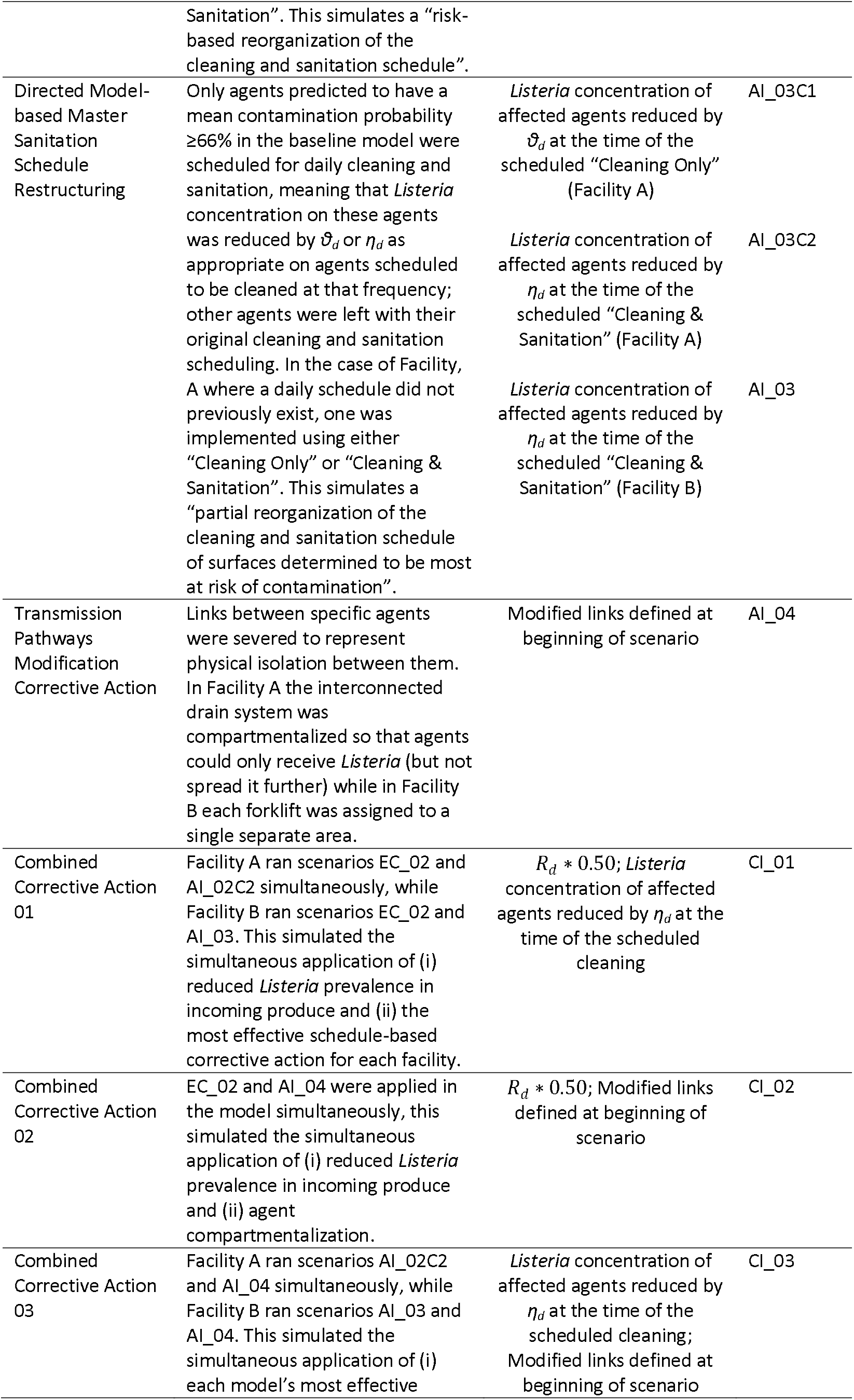
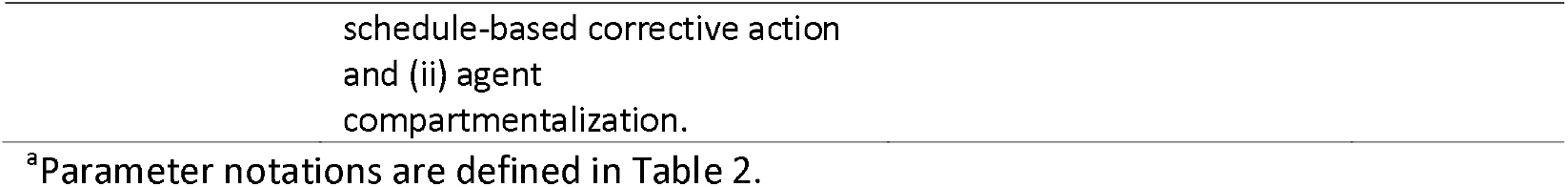
Corrective Action Scenarios and their virtual implementation within the Agent-Based Models.

Each corrective action scenario (54 in total: Facility A: 28, Facility B: 26) was evaluated by running 1,000 iterations. Scenario analyses simulations used the same fixed seed as in the baseline model to assure a fair (counterfactual) comparison among scenarios and between each scenario and the baseline model. Efficacy of a corrective action was evaluated by comparing the prevalence of contaminated agents, separately for wet and dry area, in counterfactual iterations of the baseline model and the model with a corrective action implemented using Eq. 2. The efficacy for each iteration was defined as:

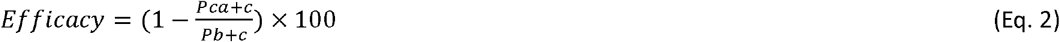

where *Pb* stands for prevalence in the baseline model iteration and *Pca* stands for prevalence in the corresponding iteration of the model with a corrective action. Constant *c* is a correction factor corresponding to 0.5/m (where *m* is the total number of agents of the area type in the model, i.e., Facility A: dry agents=171, wet agents=53; Facility B: dry agents=176 and wet agents=49); this correction factor was applied to be able to calculate the efficacy in iterations where prevalence in the baseline model was zero. Efficacy over all iterations was summarized with the median and interquartile range (IQR) statistics. The IQR was specifically used due to it being a robust measure of dispersion as it represents the middle 50% of the sample and is thus not influenced by outliers (26).

The estimate of efficacy of a corrective action provides useful information about the relative change in prevalence of contamination between the compared scenario and the baseline but it does not assess the magnitude of contamination for each scenario. Thus, the distribution of both predicted prevalence of contamination and concentration of *Listeria* on contaminated agents over all iterations were compared between the baseline and promising corrective action scenarios (identified based on sensitivity and efficacy analysis). The comparisons were presented graphically as boxplots for each scenario by wet and dry areas. For ease of interpretation, we estimated the difference between the median prevalence (expressed as percentage point (pp) difference) and between median concentration (expressed as log_10_ CFU/cm^2^) for the corrective action scenario and the baseline. Data files and code used to build the two agent-based models using NetLogo, as well as data files and R code relevant to the scenario analysis and sensitivity analysis, are available on the GitHub repository: https://github.com/IvanekLab/CPS_ABM. No statistical tests were performed to compare simulation predictions to other simulation predictions (e.g., corrective actions versus the baseline) as the modeler controls the number of replications produced (effectively “sample size”) by selecting the number of iterations and thus with sufficient computer time, there is no limit to how small a p-value value can be obtained (27). Furthermore, no comparisons were made in corrective action performance between the two Facilities (e.g., EC_04 in Facility A versus Facility B) due to key differences in their layout, schedules and specific equipment used, because of which the facility specific models were developed as opposed to a generic model; thus, statistical comparisons between the two models may lead to misleading conclusions.

## Results

### Validation

The baseline models were validated with historical data for Facilities A and B at both whole-model and area-specific levels (Fig 2; Table S10 in S1 File). All comparisons indicated lack of statistically significant differences (either with Chi Squared Test or Fisher’s Exact Test as appropriate) between agent contamination prevalence observed in historical data and mean prevalence obtained with simulated environmental sampling.

### Predicted *Listeria* prevalence and concentration in wet versus dry areas

The Facility A wet areas had median agent contamination prevalence of 25.9% and 23.1% within the Loading area and Cleaning area (i.e., the model’s “wet” area), respectively (Fig 3.A), while the Facility B Loading area and Cleaning area had respective median prevalence of 42.9% and 17.9% (Fig 3.B). “Dry” areas (i.e., combination of remaining facility areas not in proximity to water) had a lower prevalence of *Listeria* positive agents, with Facility A having medians of 6.3%, 4.1%, 1.7%, 11.1% and 7.7% for the Sorting, Tray Packing, Bag Packing, Reject and Other areas, respectively (Fig 3.A). Facility B did not feature an active Bag Packing area, but all its remaining dry areas except the Reject area had medians of 0%; the Reject area median prevalence was 21.7% (Fig 3.B). Within each area, the concentration of *Listeria* on contaminated agents was recorded and analyzed. Figs 3.C and 3.D show the low concentrations (in log_10_ CFU/cm^2^) for Facilities A and B respectively, across all wet and dry areas in the modeled facilities. Each area group contained a combination of agents belonging to Zone 1-3. Visual evaluation of agents grouped by hygienic Zone revealed an overall higher prevalence and concentration in Zone 3 agents compared to Zones 1 and 2 (Fig S2 in S1 file).

**Fig 3.**
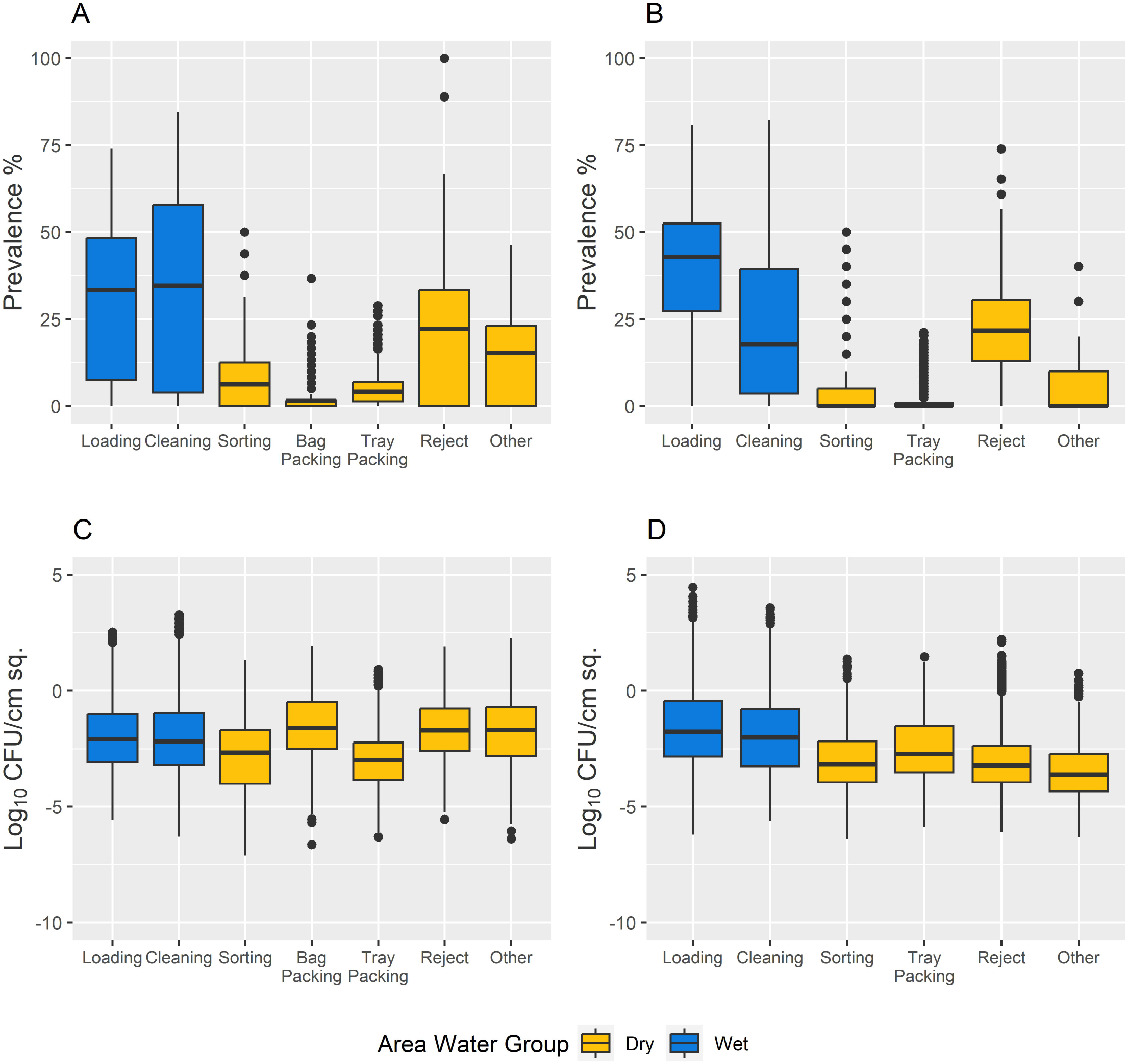
Boxplots describing *Listeria* contamination prevalence and concentration on contaminated agents on Wednesday at Midday for Facility A and B baseline conditions. Prevalence of *Listeria* contamination within each area of Facility A (panel A) and Facility B (panel B). Both facilities show higher prevalence in wet areas (blue) than dry areas (yellow), except for the Reject area. Log_10_ concentrations (CFU/cm^2^) of *Listeria* on contaminated agents within each area of Facility A (panel C) and Facility B (panel D) with median concentrations listed showing low level of contamination.

### Sensitivity Analysis

The effects of model input parameters on *P_W*, analyzed using PRCC, are depicted in Fig 4. The most influential parameters were the concentration of *Listeria* spp. per gram of contaminated raw produce (N_R_) and the probability of contact between Zone 1 agents (P_11_). In Facility A the probability of *Listeria* transfer between Zone 3 agents given contact (τ_33_) was negatively correlated with prevalence of contaminated agents, while the transfer probability between Zone 1 agents and Zone 3 agents (τ_13_) was negatively correlated in Facility B (Fig 4). In both facilities the negatively correlated parameters involved equipment that was connected to drainage systems within the facilities (as well as interconnected drains in Facility A, which are designated Zone 3); *Listeria* in the drainage system could not re-contaminate other agents and instead would show die off, which led to the negative correlation.

**Fig 4.**
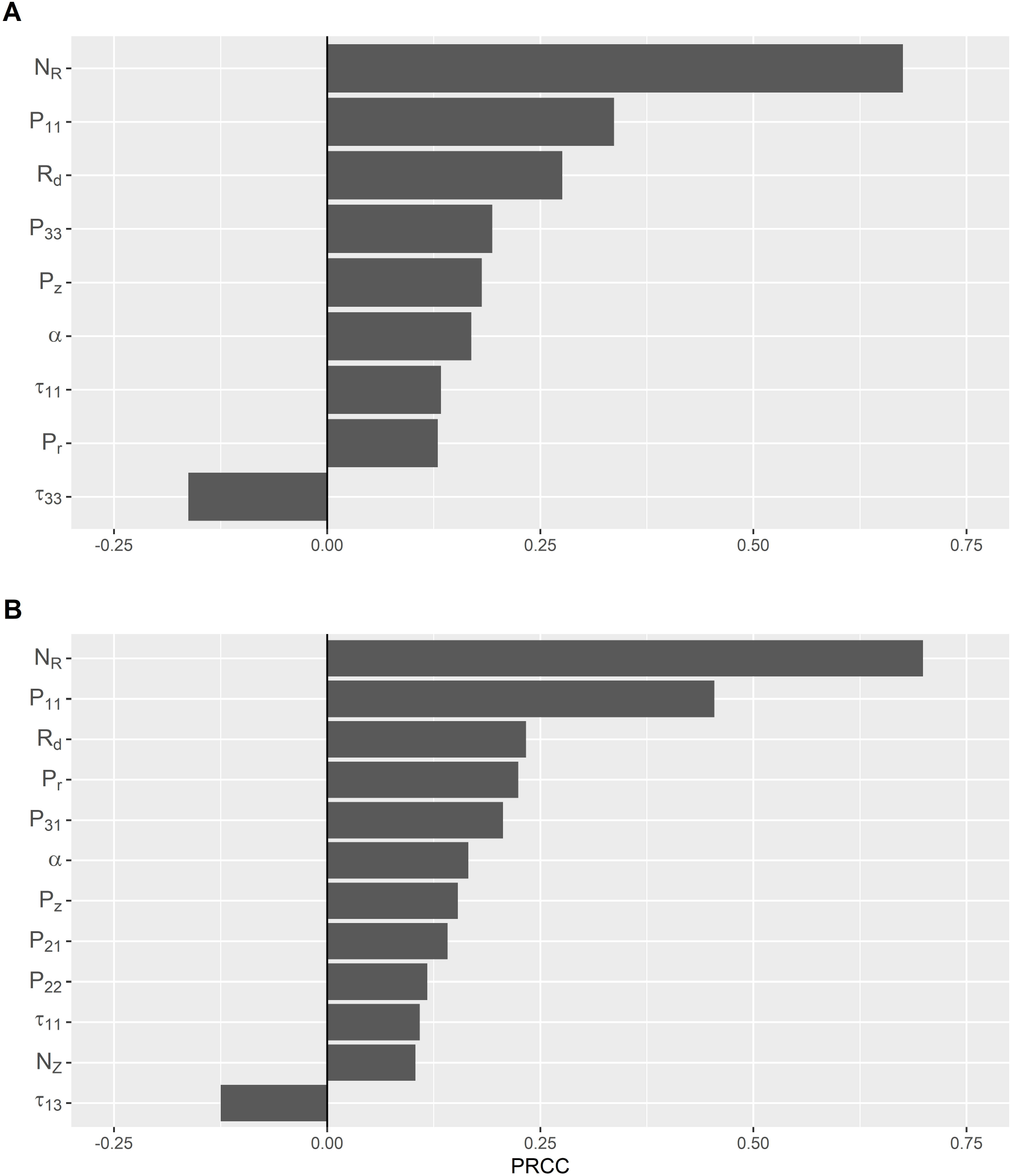
Sensitivity plots of significant model input parameters against prevalence of *Listeria* contaminated agents at midday Wednesday of the second week of simulation for each facility. Significant partial rank correlation coefficient (PRCC) values were determined using Bonferroni correction according to the number of parameters evaluated in Facilities A and B, respectively. (N_R_: Concentration of *Listeria* spp. per gram of contaminated raw produce (CFU/g); P_ij_: Probability of contact from contaminated surface in Zone *i* to another surface in Zone *j*, where *i*=*j*=Zone1, Zone 2, Zone 3 or Employee agent type; P_r_: Rate of random event occurrences that introduces *Listeria* spp. from outside the room per hour; P_z_: Probability that *Listeria* spp. is introduced into the room via objects from Zone 4 per hour; R_d_: Prevalence of *Listeria* spp. in produce on day d, for d = *Monday, Tuesday, Wednesday, Thursday, Friday*; *α*: Proportion of *Listeria* spp. transferred to a surface upon contact with a contaminated raw produce; τ_ij_: Probability of *Listeria* spp. transfer from *i* to *j* agent given contact, where *i*=*j*=Zone1, Zone 2, Zone 3, or Employee agent type.; *N*_*z*_: Amount of *Listeria* spp. introduced to an agent or patch from Zone 4 (CFU) per occurrence)

### Scenario Analysis

Comparison of modeled corrective actions with the baseline model allowed for evaluation of the efficacy of each corrective action and provided data for prioritizing strategies for virtual implementation to evaluate the magnitude of contamination for each scenario (Fig 5-6; Table S11 in S1 File). Based on efficacies that did not produce a positive result in both wet and dry areas of respective facility and sensitivity analysis results, Random Event Occurrence Reduction (PR), Random Load Reduction (LR), Z4 Event Occurrence Reduction (PZ) and Z4 Load Reduction (LZ) corrective actions were deemed ineffective (Fig S3-S6 in S1 File) and were excluded from further analysis of the magnitude of contamination.

**Fig 5.**
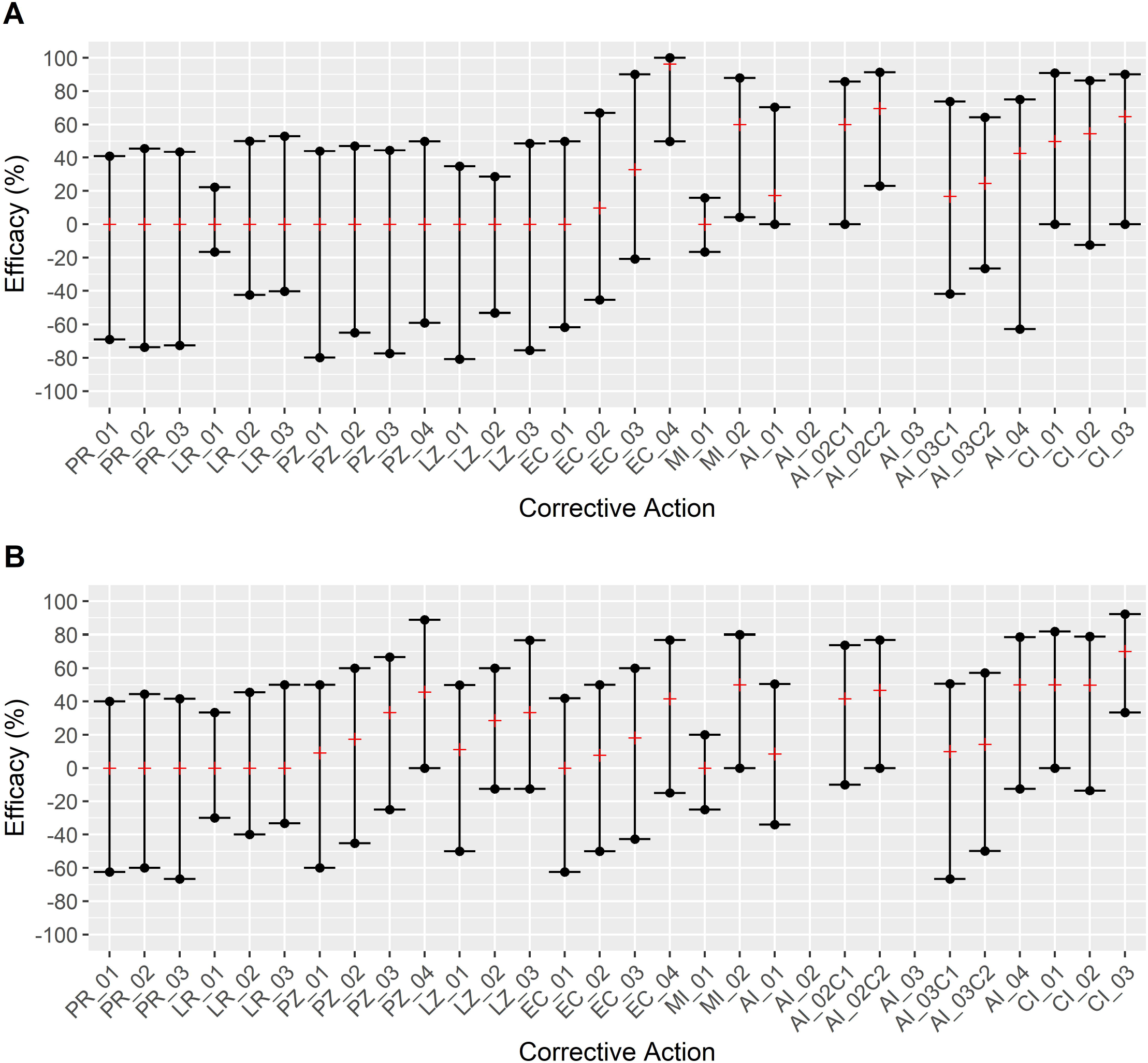
Comparison of corrective action efficacy against baseline conditions in Facility A. Efficacy was calculated using eq. 2 for each area (i.e., “wet” and “dry”) within a model for each applicable corrective action and displayed by median efficacy (red line marker) and interquartile range (black dot crossed by line marker). Positive efficacy indicated a lower *Listeria* prevalence in the model with a corrective action compared to the baseline model and thus effectiveness of the corrective action, while zero or negative efficacy indicated that the corrective action is predicted to not be able to reduce the agent contamination prevalence. Panel A: Facility A Wet area. Panel B: Facility A Dry area. PR_01-PR_03: Random Event Occurrence Reduction (125%; 150%; 175% event delay from baseline respectively). LR_01-LR_03: Random Load Reduction (1-3 Log_10_, respectively). PZ_01-PZ_04: Z4 Event Occurrence Reduction (25%; 50%; 75%; 100% reduction from baseline respectively). LZ_01-LZ_03: Z4 Load Reduction (1-3 Log_10_, respectively). EC_01-EC_04: Reduction of *Listeria* Prevalence in incoming produce (25%; 50%; 75%; 100% reduction from baseline, respectively). MI_01: Cleaning Effectiveness Improvement (increased *Listeria* removed during reduction events increased by 3 log_10_). MI_02: Weekend Deep Clean (Removal of *Listeria* from all agents regardless of cleanability status every Sunday). AI_01: Enhanced Flume Water Treatment (2 log_10_ removal of *Listeria* in flume agent per hour of production). AI_02/AI_02C1/AI_02C2: Broad Model-based Master Sanitization Restructuring (Agent cleaning and sanitation schedules were fully reassigned according to a mean contamination probability; Facility A was given a daily schedule for both “Cleaning Only” and “Cleaning & Sanitation” respectively). AI_03/AI_03C1/AI_03C2: Directed Model-based Master Sanitization Schedule Restructuring (Sanitization of agents with a mean contamination probability ≥66% was set to a daily frequency; Facility A was given a daily schedule for both “Cleaning Only” and “Cleaning & Sanitation” respectively). AI_04: Transmission Pathways Modification Corrective Action (Drain compartmentalization). CI_01: EC_02 and AI_02C2 were applied simultaneously. CI_02: EC_02 and AI_04 were applied simultaneously. CI_03: AI_02C2 and AI_04 were applied simultaneously.

**Fig 6.**
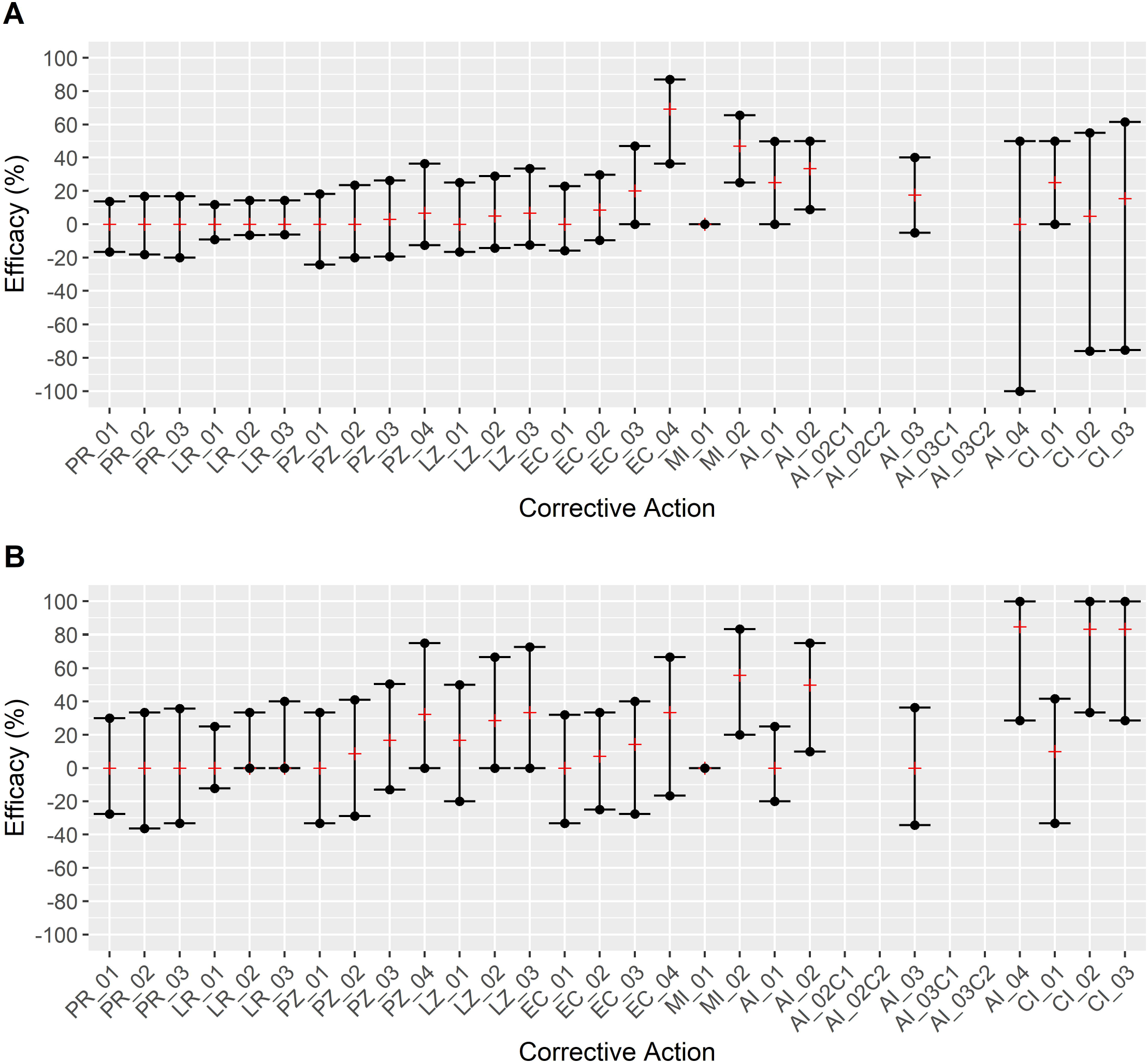
Comparison of corrective action efficacy against baseline conditions in Facility B. Efficacy was calculated using eq. 2 for each area (i.e., “wet” and “dry”) within a model for each applicable corrective action and displayed by median efficacy and interquartile range. Positive efficacy indicated a lower *Listeria* prevalence in the model with a corrective action compared to the baseline model and thus effectiveness of the corrective action, while zero or negative efficacy indicated that the corrective action is predicted to not be able to reduce the agent contamination prevalence. Panel A: Facility B Wet area. Panel B: Facility B Dry area. PR_01-PR_03: Random Event Occurrence Reduction (125%; 150%; 175% event delay from baseline respectively). LR_01-LR_03: Random Load Reduction (1-3 Log_10_, respectively). PZ_01-PZ_04: Z4 Event Occurrence Reduction (25%; 50%; 75%; 100% reduction from baseline respectively). LZ_01-LZ_03: Z4 Load Reduction (1-3 Log_10_, respectively). EC_01-EC_04: Reduction of *Listeria* Prevalence in incoming produce (25%; 50%; 75%; 100% reduction from baseline, respectively). MI_01: Cleaning Effectiveness Improvement (increased *Listeria* removed during reduction events increased by 3 log_10_). MI_02: Weekend Deep Clean (Removal of *Listeria* from all agents regardless of cleanability status every Sunday). AI_01: Enhanced Flume Water Treatment (2 log_10_ removal of *Listeria* in flume agent per hour of production). AI_02/AI_02C1/AI_02C2: Broad Model-based Master Sanitization Restructuring (Agent cleaning and sanitation schedules were fully reassigned according to a mean contamination probability). AI_03/AI_03C1/AI_03C2: Directed Model-based Master Sanitization Schedule Restructuring (Sanitization of agents with a mean contamination probability ≥66% was set to a daily frequency). AI_04: Transmission Pathways Modification Corrective Action (Separation of forklift area assignment). CI_01: EC_02 and AI_03 were applied simultaneously. CI_02: EC_02 and AI_04 were applied simultaneously. CI_03: AI_03 and AI_04 were applied simultaneously.

### Modifying Prevalence of *Listeria* Contamination in Incoming Produce

Reducing the prevalence of *Listeria* on incoming raw produce (scenarios EC_01, EC_02, EC_03 and EC_04) showed a corresponding drop in median prevalence of contamination on agents in both wet and dry areas. Facility A showed a maximum reduction (at EC_04) of 32.07 pp and 2.34 pp (Fig S7.A) for wet and dry area, respectively. Median *Listeria* concentrations on positive agents per area decreased by 0.57 log_10_ CFU/cm^2^ and 0.52 log_10_ CFU/cm^2^, respectively (Fig S7.C).

The maximum impact of reducing contamination prevalence on incoming raw produce in Facility B led to a reduction of 20.41 pp in median contamination prevalence for the wet area, and a median prevalence reduction of 1.70 pp in the dry area (Fig S7.B). Wet area *Listeria* concentrations on positive agents also showed a substantial decrease in predicted median (1.05 log_10_ CFU/cm^2^; Fig S7.D), while predicted median *Listeria* concentrations in dry areas only were reduced by 0.20 log_10_ CFU/cm^2^.

### Enhanced Cleaning and Sanitation Strategies

Improving the effectiveness of *Listeria* removal actions (“Cleaning Only” and “Cleaning & Sanitation”; scenario MI_01) by 3 log_10_ showed no meaningful changes in Facility A or Facility B’s median *Listeria* prevalence or concentration for either wet or dry areas. Weekend deep cleaning (scenario MI_02) however, led to considerable reduction of median prevalence in both models. Facility A wet area prevalence decreased by 24.53 pp (Fig S8.A) and Facility B wet area prevalence decreased by 16.33 pp (Fig S8.B).

### Agent-Targeted Corrective Actions

In Facility A the enhanced flume water treatment (scenario AI_01) and risk-based (scenarios AI_02C1 and AI_02C2) “Cleaning Only”/ “Cleaning & Sanitation” corrective actions showed the best performance in reducing both facility-wide median prevalence of contaminated agents; risk-based corrective actions are activities that target agents with higher probabilities of becoming contaminated according to the baseline model. Of the risk-based corrective actions applied, AI_02C2 had the largest impact in the wet areas (Fig S9.A), producing a median decrease in prevalence of contaminated agents against the baseline of 28.30 pp and a median decrease in concentration of 0.36 log_10_ CFU/cm^2^. In contrast, AI_01 was the most effective in Facility B’s wet area, but AI_03 was more effective in the dry area, producing a median prevalence decrease of 12.24 pp and 0.57 pp for each area respectively in AI_01, and 10.20 pp and 2.27 pp in AI_03 (Fig S9.B). Median concentration on positive agents decreased by 0.40 log_10_ CFU/cm^2^ and 0.05 log_10_ CFU/cm^2^ of each respective area in AI_01, and decreased by 0.25 log_10_ CFU/cm^2^ and 0.12 log_10_ CFU/cm^2^ for each respective area in AI_03 (Fig S9.D).

The “Transmission Pathways Modification Corrective Action” (AI_04) was applied to each model by eliminating connection links chosen to have minimal impact to facility function and a higher likelihood of implementation by a facility in response to a contamination event. This compartmentalization seeks to limit the spread of *Listeria* moving between zones (i.e., non-FCS-to-FCS transmission) by reducing the number of connections for strategically chosen agents. For example, in Facility A, the network of indoor square and trench drains was remodeled as isolated agents to simulate the introduction of anti-backflow valves within the system. This intervention reduced the Facility A wet area *Listeria* prevalence by 15.09 pp, concentration by 0.13 CFU/cm^2^, dry area prevalence by 2.92 pp and the concentration by 0.70 log_10_ CFU/cm^2^. In Facility B, the AI_04 corrective action involved assigning a single forklift to the Loading area operations, and one to its Reject area, rather than allowing both forklifts to interact with both areas. While this change in traffic patterns (i.e., connections) was effective for reducing the likelihood of *Listeria* contamination in the Facility B dry area, producing a median prevalence reduction of 3.41 pp, the amount of contamination on contaminated agents slightly increased by 0.20 log_10_ CFU/cm^2^. As the corrective action was tailored to facility, it is unsurprising that the results of AI_04 differed between models. Compartmentalizing Facility A’s drains prevented spread between wet and dry areas of the model, thus producing a facility-wide reduction in prevalence and concentration (Fig S9.A), while isolating Facility B’s forklifts only impacted prevalence in dry areas.

### Combined Corrective Actions

Of the three combined corrective actions, combinations CI_01 and CI_03 were most effective in reducing *Listeria* prevalence in Facility A wet areas (Fig S10.A; reduction of 24.53 pp for both); combination CI_01 decreased median concentration in the wet area by 0.44 log_10_ CFU/cm^2^. For the dry areas, Scenario CI_03 showed the greatest reduction and efficacy with a median concentration decrease of 0.59 log_10_ CFU/cm^2^ in the dry area (Fig S10.C).

In Facility B wet areas, CI_01 was the most effective correction action with a prevalence reduction of 14.29 pp (Fig S10.B). Median concentration among contaminated wet areas dropped by 0.39 log_10_ CFU/cm^2^ (Fig S10.D). CI_02 and CI_03 were more effective in the dry areas with a decrease in prevalence of 3.41 pp for both; median concentration among contaminated dry areas increased by 0.08 and 0.20 log_10_ CFU/cm^2^, respectively (Fig S10.D).

## Discussion

This study described the development of two ABMs of *Listeria* contamination dynamics in produce packinghouses, demonstrating their successful validation and characterization of baseline behaviors. Both facilities were functionally similar, receiving raw produce that is subsequently washed, sorted, and packed. However, the facilities differed in layout and specific food safety practices (e.g., frequency of “Cleaning Only” and “Cleaning & Sanitation” cleaning operations), which are important to consider when evaluating the risk of environmental *Listeria* contamination and mitigation strategies. Using the developed models, a range of corrective actions were tested, further demonstrating strengths of such ABMs as a decision support tool for industries. The most effective corrective actions in both models were: (i) reducing incoming *Listeria* on contaminated produce, (ii) simulation-informed modification of cleaning and sanitation strategies and (iii) eliminating specific agent-to-agent transmission pathways. While most of these corrective actions were more effective in the wet area of the respective facility than dry, eliminating specific transmission pathways (i.e., AI_04) was more effective in reducing the prevalence of Facility B’s dry area.

### *Listeria* Dynamics in Modelled Produce Packinghouses

Both models predicted elevated *Listeria* contamination within areas characterized as often containing a high level of water, such as those areas involved in raw produce loading or cleaning operations. This predicted pattern of increased prevalence in areas containing high levels of water agrees with findings regarding the water activity required for *L. monocytogenes* growth by Pietrysiak et al. and Farber et al. (28,29). However, the Reject area (which was classified as “dry”) was also predicted to show elevated *Listeria* contamination prevalence compared to other Dry areas. While there may not have been water directly involved in this Reject area, wet or damaged produce was stored in large crates in this area for extended periods. This area contains “sink” sites that receive *Listeria*, but do not transfer it to another agent (sink sites were also described by Malley et al. (30)). Importantly, increased *Listeria* prevalence does not always rely on growth, it can simply reflect increased introduction without actual growth. The predicted concentration of *Listeria* on positive agents was highly variable within all production areas in the two facilities though concentrations appeared lower in the dry areas in facility B and some of the production areas in facility A. The median predicted concentrations were relatively low in both models. It is possible that the low concentration of *Listeria* within both models reduced the probability of cross-contamination occurring, and subsequently reducing how far *Listeria* can be transmitted throughout a facility. The low concentrations on positive agents would in reality also be more difficult to detect (31).

Dry areas showed lower contamination prevalence, but their closer proximity to the end of the product line presents a risk of contaminating finished produce. These areas are less likely to be contaminated by *Listeria* on incoming raw produce due to their distance from the loading area and lack of water in the area. This does not prevent them from being contaminated through alternative means (i.e., Zone 4 introduction) based on the facility’s design. In this case, it is possible for other *Listeria* contamination routes to bypass other stages of a product line and reach finished produce more quickly. However, in these two models no Zone 2 or 3 surfaces were close enough to a Zone 4 introduction site to cause a large amount of *Listeria* contamination from it. Ultimately this is a facility-specific issue dependent on local layout and not mutually exclusive from introduction on contaminated raw produce.

Sensitivity analysis identified the concentration of *Listeria* spp. per gram of contaminated raw produce (N_R_) and probability of contact from a contaminated surface in Zone 1 to another surface in Zone 1 (P_11_) as the two most influential factors in prevalence of *Listeria* contaminated agents in both facilities. Similar to previous reports (32,33), this suggests that if incoming produce is a primary source of *Listeria* introduced to the facilities, it would be able to rapidly spread among FCSs as mediated by P_11_ and subsequently contaminate finished product. In reality, it is difficult to trace the movement of *Listeria* spp. through a facility to such a fine degree. However, it is possible that even with a low contamination prevalence a sufficiently high volume of incoming raw produce may lead to introduction of an amount of *Listeria* that is likely to spread into the rest of the facility from the initial introduction site. Berrang et al., (34) suggest a mechanism of *Listeria spp*. introduction like this may also occur in poultry processing plants. These results, combined with the lack of zone 1 data for model validation, emphasize the need for further research into postharvest *Listeria* levels on raw produce. This is both to better understand *Listeria* introduction behavior via a potentially major introduction route, and to produce more accurate models that act on *Listeria* introduced on raw contaminated produce.

### Limiting *Listeria* Introduction into Produce Packinghouses

Corrective actions applied to the models were initially designed around targeting the three routes of *Listeria* introduction into a facility (i.e., contaminated raw produce, Zone 4, or random occurrences) to assess the effectiveness of preventing “exterior” *Listeria* entering the facility. Factors to consider in these corrective actions include: facility layout (especially agent proximity to potential Zone 4 introduction sites (34,35)), postharvest contamination status of raw produce, and employee movement patterns. Incoming raw produce (the primary route in both models) has also been identified as a key vehicle of introduction in other studies involving *L. monocytogenes* (34,36–38), reinforcing the findings of both the sensitivity analysis and route-based corrective action comparisons. Although controlling the prevalence of *Listeria* on incoming raw produce can be difficult given the abundance and randomness of external sources that can contaminate produce before reaching a packinghouse (39,40), stringent implementation and verification of Good Agricultural Practices (GAP) and other supply chain programs could represent one strategy to reduce *Listeria* prevalence on incoming raw produce.

Corrective actions involving remaining introduction routes (Zone 4 or random occurrences) were eliminated early on due to poor performance in both models. In the case of Zone 4-based corrective actions some improved performance was seen in each model’s Dry area contamination prevalence (i.e., those closer to Zone 4 introduction sites), but was not effective across both areas. However, this reinforces importance of considering the facility specific layout in designing corrective actions because this type of corrective action would be more effective in facilities that have Zone 4 occurrences affecting a larger number of surfaces.

### Modifying Surface Cleanability and *Listeria* Harborage Capabilities

A second series of corrective actions were formed under the assumption that introduction could not be reduced, effectively targeting *Listeria* that has successfully entered the virtual facility. These measures involved enhancing or reorganizing existing measures used to reduce *Listeria*, such as increasing the effectiveness and frequency of sanitation events or changing in plant transmission routes (e.g., by restricting equipment such as forklifts to a specific room). While the exact implementation differed between modeled facilities, similar corrective actions have been historically implemented to control *Listeria* spread within food facilities, specifically in the forms of increased cleaning and sanitation frequency or replacing equipment with easier to clean versions (increasing cleanability), or by modifying equipment to eliminate niches (reducing harborage) (6) (both of which were implicitly and simplistically represented in the developed ABMs in corrective action scenarios that altered an agent’s cleanability, from uncleanable to cleanable).

The two types of cleaning and sanitation schedule restructuring evaluated were: (i) broad, where surfaces are eligible for cleaning and sanitation, with the mean contamination probability determining specific schedule frequency, and (ii) directed, where surfaces with a mean contamination probability ≥66% are cleaned and sanitized every day of the work week. A key difference in the *Listeria* contamination prevalence outcomes following restructuring of the cleaning and sanitation schedule between both facilities is due to differences in their initial schedules; with Facility A only performing a weekly cleaning and sanitation operation, and Facility B cleaning and sanitation at a daily frequency. As a result, introducing a higher frequency of cleaning and sanitation operations for more surfaces showed an improvement for Facility A regardless of whether the cleaning and sanitation scenario implemented is more (AI_02C2: “Cleaning & Sanitation”) or less (AI_02C1: “Cleaning Only”) comprehensive. This improvement was less effective in directed restructuring (AI_03C1/AI_03C2), as fewer surfaces were scheduled to undergo daily cleaning or cleaning and sanitation regardless of the method used. Conversely, in Facility B’s wet areas the corrective actions AI_02 and AI_03 were effective to similar degrees (having the same median prevalence in wet areas). Similarly, in Facility B’s dry areas, broad (AI_02) and directed (AI_03) rescheduling also produced similar reductions in median *Listeria* contamination prevalence (though differed from each other by a near negligible amount). These reductions in contamination prevalence in both areas of Facility B are likely due to the new cleaning and sanitization schedule in corrective actions AI_02 and AI_03 now targeting key agents in Facility B that were previously not cleaned and sanitized at a sufficient frequency in the baseline scenario. These specific agents were targeted in both corrective actions (AI_02 and AI_03) of Facility B due to their mean contamination probability being ≥66%. However, while both forms of rescheduling were effective across both areas in Facility B, it is important to reiterate that directed rescheduling (AI_03C1/AI_03C2) was not as effective as broad rescheduling (AI_02C1/AI_02C2) in Facility A. This highlights an important point that while the predicted contamination probability of an agent (i.e., a surface that the agent represents) can be a useful decision support component, it should not necessarily be the sole defining factor for determining the frequency of its cleaning and sanitation. Instead, creating cleaning and sanitation schedules should consider prior conditions and surface-specific information (such as Zone, proximity to water and connectivity) to avoid deprioritizing key equipment surfaces. Tompkin (6) showed that the exact response to detecting contamination by industry can differ by the facility, the food being handled, and equipment in question, but corrective actions typically will take into account various factors (such as existing cleaning and sanitation frequency, material composition for cleanability and harborage risks and other relevant practices). In both models some of the most effective corrective actions similarly required situation-specific interpretation and analysis to be implemented; this methodology is reinforced by the number of corrective actions built with site-specific considerations detailed by Tompkin (6).

Several corrective action scenarios evaluated here (i.e., AI_02/AI_02C1/ AI_02C2 and AI_03/AI_03C1/AI_03C2) required information on the agent-specific contamination probability. This demonstrates a distinct advantage in ABM usage in providing supplementary *in silico* data to an EMP, as the data, such as a surface’s predicted risk of being contaminated, can allow a facility to investigate and focus efforts on locations of higher predicted contamination risk. This may be of particular use in the event of a data scarcity in an EMP, which may be caused by insufficient coverage that cannot reliably detect *Listeria spp*. presence throughout a facility (41). Though a lack of data can be alleviated with intensive validation sampling (42), an ABM may be a highly practical tool in directing efforts more quickly and efficiently, ultimately saving time and money (12). Furthermore, given the practical impossibility of a facility to assess multiple corrective actions in reality, an ABM can evaluate various corrective action scenarios and advise which are more likely to be useful.

### Limiting *Listeria* Transmission Across Equipment Surfaces

Lastly, the third corrective action strategy, modifying existing surface transmission pathways, was functionally the most unique, as it wholly depended on the facility’s preexisting layout and equipment structure. While it is relatively straightforward to take a single piece of equipment in isolation and determine potential risks during production, the interaction effects between multiple surfaces may be more difficult to assess. Good Manufacturing Practices (GMPs) (43) generally require compartmentalization within a facility to limit pathogen transmission (typically referred to as hygienic zoning) but employing an ABM can allow for a more extensive review of such control strategies in a relatively rapid timeframe. Though some transmission pathways cannot be removed or modified due to their critical functions (i.e., major belts or the flume system), there are several auxiliary equipment surfaces that may present a greater risk than initially considered due to elevated connectivity between them. These transmission pathways may allow for cross-contamination outside of typical FCS-to-FCS routes.

Transmission pathways were identified in both models, that were not critical to packinghouse operations but still allowed for *Listeria* movement between areas; subsequently they were modified to restrict transmission. The layout of Facility A’s production line was predominantly designed as a one-way flow, but it featured a highly interconnected drainage system spanning multiple areas. In corrective action AI_04, the drain system in Facility A was modified to restrict drain cross-contamination, which was particularly effective in areas needing more drainage (i.e., wet areas). In practice, redesigning a facility’s entire drainage system with anti-backflow valves would take time and effort to complete, and may be more practical when constructing a new packinghouse facility. Implementing AI_04 in Facility B was more straightforward, as the activity of two forklift agents operating between two interior areas allowed for frequent *Listeria* cross-contamination between the Reject and Loading areas. Limiting a single forklift to each area severed any direct contamination routes, leaving *Listeria* only able to follow the production line to reach the Reject area. This compartmentalization directly limited the spread of *Listeria* into other areas and demonstrates that a relatively simple corrective action can have widespread impact. In practice, reducing surface interconnectivity, may also be implemented through employee training or redesign of equipment to reduce or prevent cross-contamination (44).

### Combined Corrective Actions Have Facility-Wide Impact on *Listeria* Harborage

As stated previously, a major advantage of an ABM is the ability to generate predictions specific to individual equipment surfaces and for specific simulated corrective actions, or their combinations, and subsequently direct efforts in more focused course. For both models, a useful metric of measuring the performance of a corrective action was investigating its efficacy and comparing change in the *Listeria* contamination prevalence in wet and dry areas. The three combined corrective action scenarios (CI_01-CI_03) were selected from individual corrective action types that showed the highest *Listeria* contamination prevalence reductions in either area: reducing the *Listeria* prevalence on incoming raw produce, cleaning and sanitation schedule restructuring and transmission pathway modification. Of these options, a 50% reduction in *Listeria* prevalence on raw produce was chosen as a more plausible outcome than complete elimination of contamination on incoming raw produce, and each facility had its own best-performing respective cleaning and sanitation restructuring option chosen. These combined corrective actions could then be simulated and further analyzed themselves to determine the performance of using multiple corrective actions simultaneously. While this is largely similar to previous scenarios, combining corrective actions is an already suggested general strategy (45) and has the benefit of relying on multiple layers of defense. The selection and evaluation of which specific corrective actions to combine however, can be more systematically done with the additional performance data provided by an ABM’s simulations.

### Limitations and Future Directions

It should be noted that a fundamental limitation facing both models was the relatively small amount of historical data available (including complete absence of historical data for Zone 1 agents), limiting the extent of validation. It is also possible that low historical prevalence in certain areas in the facility may make it more difficult to detect meaningful levels of improvement, given an already low baseline to begin with. Additionally, both models were simulated on a virtual timescale of two weeks, making for relatively short-term observations (though at the same time addressing the industry needs for decision support tools for short-term planning). Various interventions may have far more noticeable consequences to their facilities if observed for a longer period, which should be subject of a future investigation. Furthermore, there was insufficient data available on individual agent attributes with respect to their composition or materials and how this may affect cleaning and sanitation operations, requiring agents to be treated uniformly in this regard. Currently the model can support this to a limited degree by setting agents to be cleanable or uncleanable. However, this model system can support the addition of information such as composition/materials and its impact on cleaning and sanitation once acquired during future modeling applications. A related cleaning limitation involved a lack of information on how likely agents were to undergo successful cleaning or cleaning & sanitization; both of which were set at the assumed value of 0.99. Sensitivity analysis and testing lower values (i.e., γ=0.95 and δ=0.85) resulted in no meaningful change in prevalence of contaminated agents or concentration on contaminated agents. It should be noted however, that the two-week duration of this model may be too short for these values to be particularly impactful. Thus, testing the effect of the probability of successful cleaning over longer simulation runs should be a subject of a future investigation to determine whether this knowledge gap should be prioritized for research. An additional limitation due to insufficient data was that incoming crates in both facilities were treated as independent from each other, rather than simulate the arrival of clusters of crates containing contaminated produce. Again, this issue may be overcome in future models with collection of more extensive data to integrate. Due to a lack of postharvest contamination data on produce, the amount of *Listeria* introduced to the model on contaminated produce was calculated using data from Chen et al., 2016 (15) which evaluated stone fruits involved in an outbreak of listeriosis. We acknowledge that values from outbreaks may be higher than what would be expected outside of outbreak situations. Lastly, both models use a non-specific virtual strain of *Listeria* spp., but we acknowledge that different strains of *L. monocytogenes* may have differences in properties, which could be included in future modelling efforts as well.

Future models should also include economic factors to assess the most appropriate interventions or combinations of interventions that should be implemented in a given facility. While the first reaction may be to implement as many separate corrections as possible for overlapping protection, each extra layer will incur additional costs (46). Instead, being able to identify potentially more cost-effective actions, such as employee training to reduce cross contamination (47) instead of equipment replacement, would allow for better decision-making that optimizes resource allocation. Furthermore, incorporating economic factors could allow for the model to estimate the overall cost to a facility of having to operate with more systemic issues, such as layout and drainage methods (48). A key takeaway that is applicable to any type of packinghouse or food processing facility attempting to combat ongoing *Listeria* contamination is that each facility should be treated uniquely and addressed with specifically designed corrective actions that have highest potential effectiveness (49). Models such as those developed here could aid design of the facility specific corrective actions.

An additional direction for future development may include the use of ABMs in testing measures already implemented within a facility under hypothetical situations of an increased contamination risk. While the scenarios demonstrated here were to trial corrective actions, similar techniques could be used to test the impact of increased *Listeria* contamination (e.g., in incoming material) on facility and finished product contamination under the currently implemented procedures in a facility. This would expand the scope that these models can be applied to, allowing them to be used in both a diagnostic capacity for solving existing contamination issues, and to assist in pre-emptively assessing how well a facility would be able to reduce *Listeria* contamination risks in the event of a system failure.

## Conclusions

Once established within a packinghouse, *Listeria* spp. has proven to be difficult to control, and decision support tools such as the ABM reported may be valuable in not only quantifying how contamination may move through a facility, but in finding effective options for combating it. With Facilities A and B, we have illustrated that ABMs can serve as highly adaptable tools in the field of food safety through their ability to replicate the unique components of individual produce packinghouses. From our ABM scenarios, targeting *Listeria* that is introduced through the primary contamination route (in this case contaminated incoming raw produce) was the most effective method in prevalence reduction, and may be generalizable to different facilities as its implementation does not depend on a facility’s specific layout. However, assessing the effectiveness of this strategy relies on contamination data that currently are rarely available. Therefore, it may be more practical to focus on designing in-house corrective actions, such as increasing the frequency at which select surfaces are sanitized and employing measures to limit contamination spread between equipment surfaces, that account for facility-wide conditions and patterns. An element of particular note in this regard is the local presence of water within an area, as it has shown to affect *Listeria* growth and the performance of corrective actions within the specific area. The *in silico* data generated by ABMs has also shown to be useful for designing and evaluating additional corrective action scenarios. Combining contamination and corrective action results from an ABM with relevant economics data would further aid in determining the overall feasibility of implementing corrective actions specifically to individual facilities.

## Data Availability

Both agent-based models, as well as their data files and the analysis code used are available on GitHub (https://github.com/IvanekLab/CPS_ABM/)

https://doi.org/10.5281/zenodo.5921808

https://github.com/IvanekLab/CPS_ABM/

## Acknowledgments

We would like to thank the personnel representing two packinghouses for their time and providing access to their facilities.

## Supporting information

**S1 File. Supporting Information File – S1.docx**.

## Notes

### Competing Interest Statement

Dr. Zoellner is presently employed by iFoodDecisionSciences, Inc., a company that provides industry with modeling and data analysis tools.
Drs. Ivanek and Wiedmann are named as co-inventors, along with Dr. Zoellner, on an algorithm that uses an in silico method to reproduce the behavior of biological contaminants in built environments, which was licensed to iFoodDecisionSciences, Inc.

### Funding Statement

R.I., 2019CPS06, Center for Produce Safety, https://doi.org/10.13039/100008678
M.W., 2020CPS10, Center for Produce Safety, https://doi.org/10.13039/100008678
R.I., USDA-AMS-TM-SCBGP-G-18-003, Florida Department of Agriculture and Consumer Services Specialty (FLDACS), https://doi.org/10.13039/100011508
I.T., 2020-67021-32855, USDA | National Institute of Food and Agriculture (NIFA), https://doi.org/10.13039/100005825
M.W., USDA-AMS-TM-SCBGP-G-19-003, Washington State Department of Agriculture (WSDA), https://doi.org/10.13039/100010251
M.W., 2019-51181-30016, USDA | National Institute of Food and Agriculture (NIFA), https://doi.org/10.13039/100005825
The funders had no role in study design, data collection and analysis, decision to publish, or preparation of the manuscript.
CZ, iFoodDecisionSciences, Inc. https://www.ifoodds.com/
The funder provided support in the form of salaries for author C.Z. but did not have any additional role in the study design, data collection and analysis, decision to publish, or preparation of the manuscript.

