## Supplemental Data 1 for "Using agent-based modeling to compare corrective actions for *Listeria* contamination in produce packinghouses"

### Supporting information

#### Parameter Calculation

Concentration of *Listeria* spp. per gram of contaminated raw produce (CFU/g) (N_R_) was calculated by converting data from Chen et al.’s (1) Table 1 into CFU/g using total CFU/fruit and the average fruit mass from recorded minimum and maximum (136g). A gamma distribution was then constructed to match calculated minimum, maximum, median, and mean average values as closely as possible.

Average log reduction (*θ_d_* and *η_d_*) was calculated by taking values from pages 42-43 of the FDA/FSIS’s 2013 interagency risk analysis of *Listeria monocytogenes* in Retail Delicatessens (2): “Following a complete literature review (3), (min, mode, max) is set to (-1, -0.5, 0) for all “wiping” processes, to (-1.5, -0.5, 0) for all “washing” processes, and to (-8, -6, -1.5) for all “washing and sanitizing” processes.”. These values were used in Pert distribution (*θd* and *ηd*) with a default peakedness parameter value of 4.

#### Model Schedules

**Table S1. Facility A baseline operations event schedule.**

| Day/Hour | 1 | 2 | 3 | 4 | 5 | 6 | 7 | 8 | 9 | 10 | 11 | 12 | 13 | 14 | 15 | 16 | 17 | 18 | 19 | 20 | 21 | 22 | 23 | 24 |
| --- | --- | --- | --- | --- | --- | --- | --- | --- | --- | --- | --- | --- | --- | --- | --- | --- | --- | --- | --- | --- | --- | --- | --- | --- |
| Sunday | EM | EM | EM | EM | EM | EM | EM | EM | EM | EM | EM | EM | EM | EM | EM | EM | EM | EM | EM | EM | EM | EM | EM | EM |
| Monday | EM | EM | EM | EM | EM | EM | CL | CL | CL | CL | CL | PO | EM | PD | PD | PD | PD | EM | EM | EM | EM | EM | EM | EM |
| Tuesday | EM | EM | EM | EM | EM | EM | EM | EM | PO | PD | PD | PD | EM | PD | PD | PD | PD | EM | EM | EM | EM | EM | EM | EM |
| Wednesday | EM | EM | EM | EM | EM | EM | EM | EM | PO | PD | PD | PD | EM | PD | PD | PD | PD | EM | EM | EM | EM | EM | EM | EM |
| Thursday | EM | EM | EM | EM | EM | EM | EM | EM | PO | PD | PD | PD | EM | PD | PD | PD | PD | EM | EM | EM | EM | EM | EM | EM |
| Friday | EM | EM | EM | EM | EM | EM | EM | EM | PO | PD | PD | PD | EM | PD | PD | PD | PD | EM | EM | EM | EM | EM | EM | EM |
| Saturday | EM | EM | EM | EM | EM | EM | CL | CL | CL | CL | CL | CL | EM | EM | EM | EM | EM | EM | EM | EM | EM | EM | EM | EM |

EM: empty, CL: cleaning, PO: pre-op, PD: production.

**Table S2. Facility B baseline operations event schedule.**

| Day/Hour | 1 | 2 | 3 | 4 | 5 | 6 | 7 | 8 | 9 | 10 | 11 | 12 | 13 | 14 | 15 | 16 | 17 | 18 | 19 | 20 | 21 | 22 | 23 | 24 |
| --- | --- | --- | --- | --- | --- | --- | --- | --- | --- | --- | --- | --- | --- | --- | --- | --- | --- | --- | --- | --- | --- | --- | --- | --- |
| Sunday | EM | EM | EM | EM | EM | EM | EM | EM | EM | EM | EM | EM | EM | EM | EM | EM | EM | EM | EM | EM | EM | EM | EM | EM |
| Monday | EM | EM | EM | EM | EM | EM | EM | EM | PO | PD | PD | PD | EM | PD | PD | PD | PD | CL | CL | EM | EM | EM | EM | EM |
| Tuesday | EM | EM | EM | EM | EM | EM | EM | EM | PO | PD | PD | PD | EM | PD | PD | PD | PD | CL | CL | EM | EM | EM | EM | EM |
| Wednesday | EM | EM | EM | EM | EM | EM | EM | EM | PO | PD | PD | PD | EM | PD | PD | PD | PD | CL | CL | EM | EM | EM | EM | EM |
| Thursday | EM | EM | EM | EM | EM | EM | EM | EM | PO | PD | PD | PD | EM | PD | PD | PD | PD | CL | CL | EM | EM | EM | EM | EM |
| Friday | EM | EM | EM | EM | EM | EM | EM | EM | PO | PD | PD | PD | EM | PD | PD | PD | PD | CL | CL | EM | EM | EM | EM | EM |
| Saturday | EM | EM | EM | EM | EM | EM | EM | EM | EM | EM | EM | EM | EM | EM | EM | EM | EM | CL | CL | EM | EM | EM | EM | EM |

EM: empty, CL: cleaning, PO: pre-op, PD: production.

**Table S3. Facility A operations event schedule with corrective actions that involved changes in daily cleaning schedules (AI_02C1/AI_02C2/ AI_03C1/AI_03C2).**

| Day/Hour | 1 | 2 | 3 | 4 | 5 | 6 | 7 | 8 | 9 | 10 | 11 | 12 | 13 | 14 | 15 | 16 | 17 | 18 | 19 | 20 | 21 | 22 | 23 | 24 |
| --- | --- | --- | --- | --- | --- | --- | --- | --- | --- | --- | --- | --- | --- | --- | --- | --- | --- | --- | --- | --- | --- | --- | --- | --- |
| Sunday | EM | EM | EM | EM | EM | EM | EM | EM | EM | EM | EM | EM | EM | EM | EM | EM | EM | EM | EM | EM | EM | EM | EM | EM |
| Monday | EM | EM | EM | EM | EM | EM | EM | EM | EM | EM | EM | PO | EM | PD | PD | PD | CL | CL | EM | EM | EM | EM | EM | EM |
| Tuesday | EM | EM | EM | EM | EM | EM | EM | EM | PO | PD | PD | PD | EM | PD | PD | PD | CL | CL | EM | EM | EM | EM | EM | EM |
| Wednesday | EM | EM | EM | EM | EM | EM | EM | EM | PO | PD | PD | PD | EM | PD | PD | PD | CL | CL | EM | EM | EM | EM | EM | EM |
| Thursday | EM | EM | EM | EM | EM | EM | EM | EM | PO | PD | PD | PD | EM | PD | PD | PD | CL | CL | EM | EM | EM | EM | EM | EM |
| Friday | EM | EM | EM | EM | EM | EM | EM | EM | PO | PD | PD | PD | EM | PD | PD | PD | CL | CL | EM | EM | EM | EM | EM | EM |
| Saturday | EM | EM | EM | EM | EM | EM | CL | CL | CL | CL | CL | CL | EM | EM | EM | EM | EM | EM | EM | EM | EM | EM | EM | EM |

EM: empty, CL: cleaning, PO: pre-op, PD: production.

**Table S4. Facility A operations event schedule with the Sunday deep cleaning corrective action (MI_02).**

| Day/Hour | 1 | 2 | 3 | 4 | 5 | 6 | 7 | 8 | 9 | 10 | 11 | 12 | 13 | 14 | 15 | 16 | 17 | 18 | 19 | 20 | 21 | 22 | 23 | 24 |
| --- | --- | --- | --- | --- | --- | --- | --- | --- | --- | --- | --- | --- | --- | --- | --- | --- | --- | --- | --- | --- | --- | --- | --- | --- |
| Sunday | EM | EM | EM | EM | EM | EM | EM | EM | EM | EM | EM | EM | EM | CL | CL | CL | CL | CL | EM | EM | EM | EM | EM | EM |
| Monday | EM | EM | EM | EM | EM | EM | CL | CL | CL | CL | CL | PO | EM | PD | PD | PD | EM | EM | EM | EM | EM | EM | EM | EM |
| Tuesday | EM | EM | EM | EM | EM | EM | EM | EM | PO | PD | PD | PD | EM | PD | PD | PD | EM | EM | EM | EM | EM | EM | EM | EM |
| Wednesday | EM | EM | EM | EM | EM | EM | EM | EM | PO | PD | PD | PD | EM | PD | PD | PD | EM | EM | EM | EM | EM | EM | EM | EM |
| Thursday | EM | EM | EM | EM | EM | EM | EM | EM | PO | PD | PD | PD | EM | PD | PD | PD | EM | EM | EM | EM | EM | EM | EM | EM |
| Friday | EM | EM | EM | EM | EM | EM | EM | EM | PO | PD | PD | PD | EM | PD | PD | PD | EM | EM | EM | EM | EM | EM | EM | EM |
| Saturday | EM | EM | EM | EM | EM | EM | CL | CL | CL | CL | CL | CL | EM | EM | EM | EM | EM | EM | EM | EM | EM | EM | EM | EM |

EM: empty, CL: cleaning, PO: pre-op, PD: production.

**Table S5. Facility B operations event schedule with the Sunday deep cleaning corrective action (MI_02).**

| Day/Hour | 1 | 2 | 3 | 4 | 5 | 6 | 7 | 8 | 9 | 10 | 11 | 12 | 13 | 14 | 15 | 16 | 17 | 18 | 19 | 20 | 21 | 22 | 23 | 24 |
| --- | --- | --- | --- | --- | --- | --- | --- | --- | --- | --- | --- | --- | --- | --- | --- | --- | --- | --- | --- | --- | --- | --- | --- | --- |
| Sunday | EM | EM | EM | EM | EM | EM | EM | EM | EM | EM | EM | EM | EM | CL | CL | CL | CL | CL | EM | EM | EM | EM | EM | EM |
| Monday | EM | EM | EM | EM | EM | EM | EM | EM | PO | PD | PD | PD | EM | PD | PD | PD | CL | CL | EM | EM | EM | EM | EM | EM |
| Tuesday | EM | EM | EM | EM | EM | EM | EM | EM | PO | PD | PD | PD | EM | PD | PD | PD | CL | CL | EM | EM | EM | EM | EM | EM |
| Wednesday | EM | EM | EM | EM | EM | EM | EM | EM | PO | PD | PD | PD | EM | PD | PD | PD | CL | CL | EM | EM | EM | EM | EM | EM |
| Thursday | EM | EM | EM | EM | EM | EM | EM | EM | PO | PD | PD | PD | EM | PD | PD | PD | CL | CL | EM | EM | EM | EM | EM | EM |
| Friday | EM | EM | EM | EM | EM | EM | EM | EM | PO | PD | PD | PD | EM | PD | PD | PD | CL | CL | EM | EM | EM | EM | EM | EM |
| Saturday | EM | EM | EM | EM | EM | EM | EM | EM | EM | EM | EM | EM | EM | EM | EM | EM | CL | CL | EM | EM | EM | EM | EM | EM |

EM: empty, CL: cleaning, PO: pre-op, PD: production.

#### Model Specifications

**Table S6. Probability of agent contact description, equation and distribution, summary values and sources.**

| Symbol | Description | Equation/Distribution | Mean | 5th-95th Percentile | Reference |
| --- | --- | --- | --- | --- | --- |
| P_11_ | Probability of contact from contaminated surface in Zone 1 to another surface in Zone 1 | Pert(0,0.1,0.9,4) | 0.22 | [0.03, 0.5] | (4) |
| P_12_ | Probability of contact from contaminated surface in Zone 1 to another surface in Zone 2 | Pert(0.001,0.2,0.8,4) | 0.27 | [0.06, 0.53] | (4) |
| P_13_ | Probability of contact from contaminated surface in Zone 1 to another surface in Zone 3 | Pert(0.001,0.15,0.85,4) | 0.24 | [0.04, 0.51] | (4) |
| P_14_ | Probability of contact from contaminated surface in Zone 1 to another surface in Zone 4 | Pert(0.001,0.1,0.8,4) | 0.20 | [0.03, 0.45] | (4) |
| P_21_ | Probability of contact from contaminated surface in Zone 2 to another surface in Zone 1 | Pert(0,0.2,0.95,4) | 0.29 | [0.06, 0.6] | (4) |
| P_22_ | Probability of contact from contaminated surface in Zone 2 to another surface in Zone 2 | Pert(0.00005,0.15,0.7,4) | 0.22 | [0.04, 0.44] | (4) |
| P_23_ | Probability of contact from contaminated surface in Zone 2 to another surface in Zone 3 | Pert(0.001,0.2,0.85,4) | 0.28 | [0.06, 0.55] | (4) |
| P_24_ | Probability of contact from contaminated surface in Zone 2 to another surface in Zone 4 | Pert(0.05,10,80,4) | 0.20 | [0.03, 0.45] | (4) |
| P_31_ | Probability of contact from contaminated surface in Zone 3 to another surface in Zone 1 | Pert(0,0.2,0.9,4) | 0.16 | [0.01, 0.43] | (4) |
| P_32_ | Probability of contact from contaminated surface in Zone 3 to another surface in Zone 2 | Pert(0,0.035,0.9,4) | 0.17 | [0.02, 0.44] | (4) |
| P_33_ | Probability of contact from contaminated surface in Zone 3 to another surface in Zone 3 | Pert(0.0002,0.125,0.85,4) | 0.23 | [0.04, 0.49] | (4) |
| P_34_ | Probability of contact from contaminated surface in Zone 3 to another surface in Zone 4 | Pert(0.0001,0.02,0.6,4) | 0.11 | [0.01, 0.29] | (4) |
| P_41_ | Probability of contact from contaminated surface in Zone 4 to another surface in Zone 1 | Pert(0,0.1,0.95,4) | 0.23 | [0.03, 0.52] | (4) |
| P_42_ | Probability of contact from contaminated surface in Zone 4 to another surface in Zone 2 | Pert(0.0001,0.1,0.8,4) | 0.20 | [0.03, 0.45] | (4) |
| P_43_ | Probability of contact from contaminated surface in Zone 4 to another surface in Zone 3 | Pert(0,0.1,0.9,4) | 0.22 | [0.03, 0.5] | (4) |
| P_44_ | Probability of contact from contaminated surface in Zone 4 to another surface in Zone 4 | Pert(0,0.05,0.4,4) | 0.10 | [0.01, 0.23] | (4) |

**Table S7. Transfer Coefficient (TC) matrix based on Zone or employee type^1^.**

|  | 1 | 2 | 3 | Employee (e) |
| --- | --- | --- | --- | --- |
| 1 | -1.51 | -3.68 | -0.28 | -1.97 |
| 2 | -1.51 | -1.51 | -3.53 | -1.97 |
| 3 | -0.31 | -0.31 | -0.31 | -0.82 |
| e | -1.97 | -1.97 | -1.69 | -3.43 |

^1^Transfer Coefficient was selected based on a combination of the sender (i: horizontal) and receiver (j: vertical) (3,5)

**Table S8. Standard Deviation (STD) matrix based on Zone or employee type^1^.**

|  | 1 | 2 | 3 | Employee (e) |
| --- | --- | --- | --- | --- |
| 1 | 0.2 | 0.2 | 0.2 | 0.87 |
| 2 | 0.2 | 0.2 | 0.2 | 0.87 |
| 3 | 0.2 | 0.2 | 0.2 | 0.87 |
| e | 0.87 | 0.87 | 0.87 | 0.79 |

^1^Standard Deviation was selected based on a combination of the sender (i: horizontal) and receiver (j: vertical) (3,5)

**Table S9. Mean and 5^th^-95^th^ percentiles of probability of *Listeria* spp*.* transfer between agent types given contact^1^.**

|  | 1 | 2 | 3 | Employee (e) |
| --- | --- | --- | --- | --- |
| 1 | $\begin{matrix} 3.44E-02 \\ \left[ 1.45E-02, 6.59E-02 \right] \end{matrix}$ | $\begin{matrix} 3.44E-02 \\ \left[ 1.45E-02, 6.59E-02 \right] \end{matrix}$ | $\begin{matrix} 5.45E-01 \\ \left[ 2.30E-01, 1.04E+00 \right] \end{matrix}$ | $\begin{matrix} 7.97E-02 \\ \left[ 3.97E-04, 2.89E-01 \right] \end{matrix}$ |
| 2 | $\begin{matrix} 2.32E-04 \\ \left[ 9.80E-05, 4.46E-04 \right] \end{matrix}$ | $\begin{matrix} 3.44E-02 \\ \left[ 1.45E-02, 6.59E-02 \right] \end{matrix}$ | $\begin{matrix} 5.45E-01 \\ \left[ 2.30E-01, 1.04E+00 \right] \end{matrix}$ | $\begin{matrix} 7.96E-02 \\ \left[ 3.97E-04, 2.89E-01 \right] \end{matrix}$ |
| 3 | $\begin{matrix} 5.83E-01 \\ \left[ 2.46E-01, 1.12E+00 \right] \end{matrix}$ | $\begin{matrix} 3.28E-04 \\ \left[ 1.38E-04, 6.29E-04 \right] \end{matrix}$ | $\begin{matrix} 5.45E-01 \\ \left[ 2.30E-01, 1.04E+00 \right] \end{matrix}$ | $\begin{matrix} 1.52E-01 \\ \left[ 7.56E-04, 5.52E-01 \right] \end{matrix}$ |
| e | $\begin{matrix} 7.98E-02 \\ \left[ 3.97E-04, 2.89E-01 \right] \end{matrix}$ | $\begin{matrix} 7.98E-02 \\ \left[ 3.97E-04, 2.89E-01 \right] \end{matrix}$ | $\begin{matrix} 1.13E+00 \\ \left[ 5.61E-03, 4.09E+00 \right] \end{matrix}$ | $\begin{matrix} 1.95E-3 \\ \left[ 1.86E-05, 7.39E-03 \right] \end{matrix}$ |

^1^Transfer coefficient (TC) (Table S7) and standard deviation (STD) (Table S8) were selected based on agent type, then calculated via ${10}^{Normal(TC,STD)}$. (3,5)

#### Validation

**Table S10. Comparison of results of Historical and Simulated Sampling and testing for *Listeria* contamination in Facilities A and B.**

|  | Historical Sampling | | | Simulated Sampling | p-value |
| --- | --- | --- | --- | --- | --- |
|  | Total | Positive | Prevalence (%, 95% CI)^a^ | Mean Prevalence (%, 5^th^ and 95^th^ percentile) |  |
| **Facility A** | | | | | |
| All Samples | 102 | 16 | 16 (10-24) | 11 (6-19) | 0.33^b^ |
| Wet Agents | 56 | 14 | 25 (16-38) | 15 (8-27) | 0.19^c^ |
| Dry Agents | 46 | 2 | 4 (1-15) | 7 (2-18) | 1.00^c^ |
| Zone 2 | 55 | 4 | 7 (3-17) | 4 (1-13) | 0.68^c^ |
| Zone 3 | 43 | 12 | 28 (17-43) | 20 (11-34) | 0.38^b^ |
| **Facility B** | | | | | |
| All Samples | 174 | 26 | 15 (10-21) | 11 (7-17) | 0.30^b^ |
| Wet Agents | 90 | 21 | 23 (16-33) | 17 (10-26) | 0.27^b^ |
| Dry Agents | 79 | 4 | 5 (2-12) | 8 (4-17) | 0.39^b^ |
| Zone 2 | 123 | 11 | 9 (5-15) | 8 (5-15) | 0.85^b^ |
| Zone 3 | 29 | 7 | 24 (12-42) | 19 (9-38) | 0.63^b^ |

^a^95% Confidence interval based on the Wilson score interval method. ^b^Chi Square Test analysis. ^c^Fisher’s Exact Test analysis.

#### Probability of Proper Agent Cleaning

Alternative values of the probability that a cleanable agent was properly cleaned when “Cleaning & Sanitation” (γ) or “Cleaning Only” (δ) was performed were tested (γ=0.95 and δ=0.85). This resulted at most in extremely small changes in both Facilities A and B’s outcomes of interest (Fig S1).

| 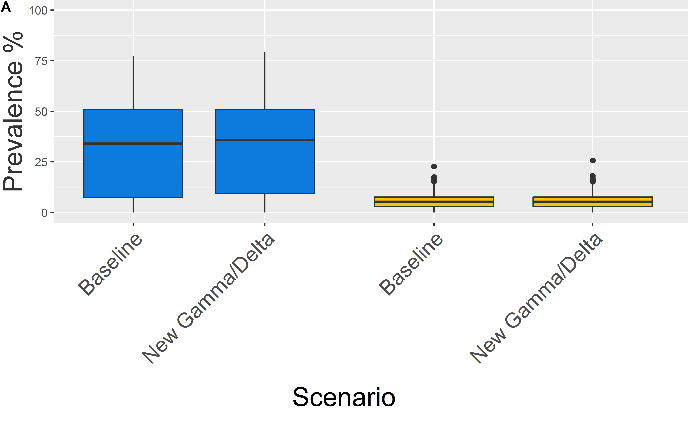 | 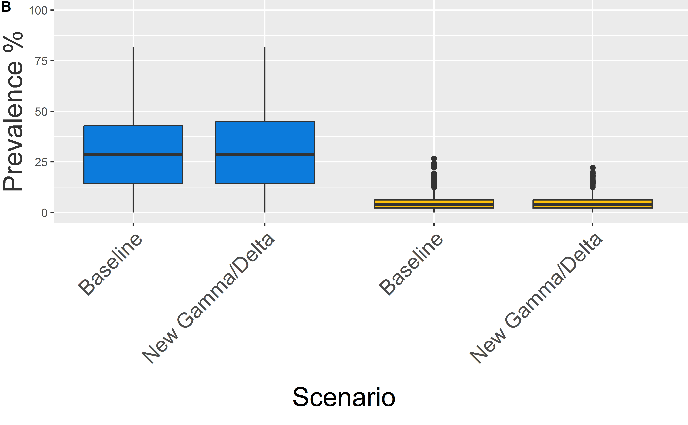 |
| --- | --- |
| 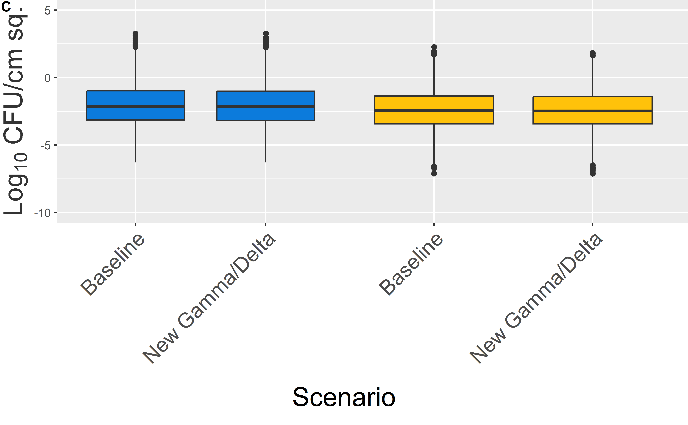 | 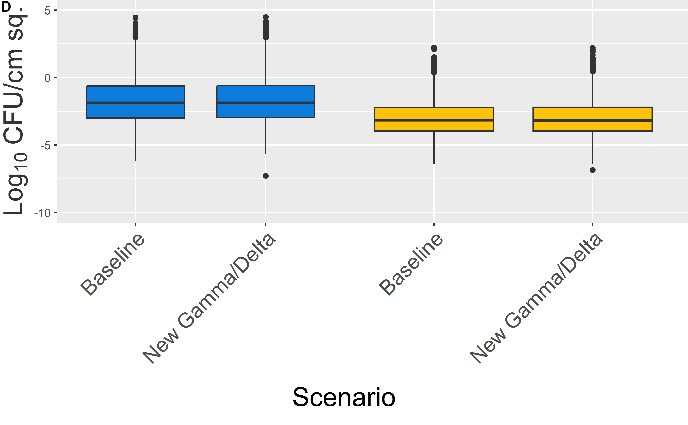 |

**Fig S1.** **Boxplots describing the prevalence and concentration on contaminated agents on Wednesday at Midday for Facility A and B on both wet (blue) and dry (yellow) area agents the baseline model (γ=0.99 and δ=0.99) and alternate values (γ=0.95 and δ=0.85).** A: Facility A *Listeria* contamination prevalence of all agents in wet and dry areas. B: Facility B *Listeria* contamination prevalence of all agents in wet and dry areas. C: Facility A *Listeria* log_10_ concentrations on all positive agents in wet and dry areas. D: Facility B *Listeria* log_10_ concentrations on all positive agents in wet and dry areas.

#### Predicted *Listeria* prevalence and concentration in Zones 1-3

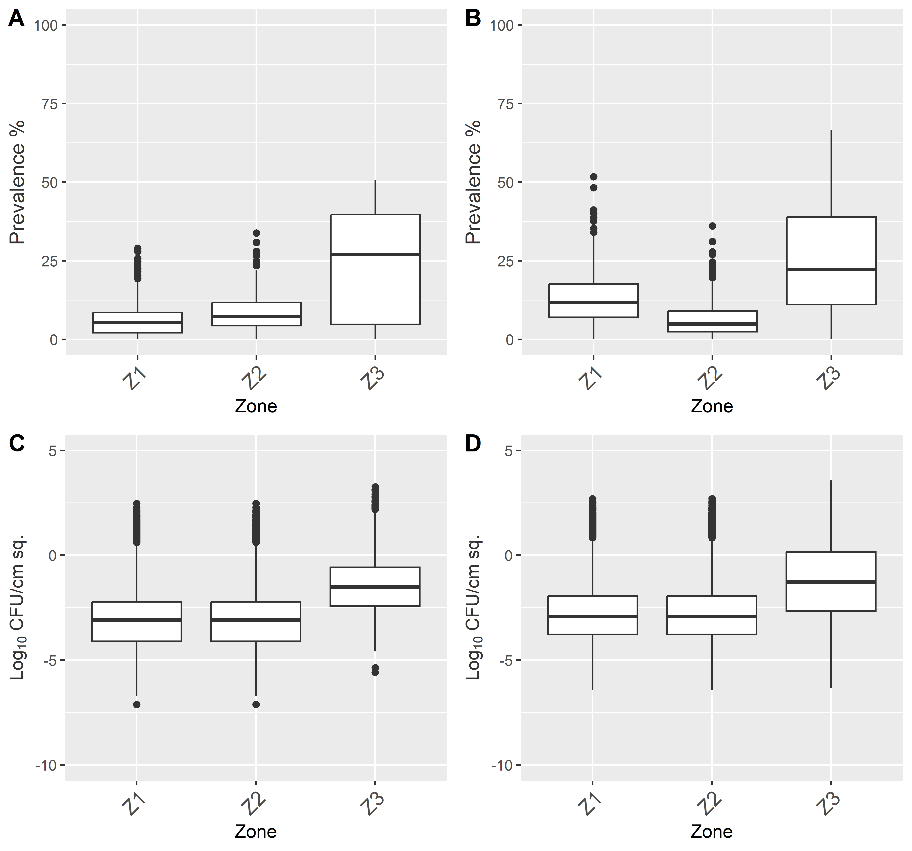

**Fig S2. Boxplots describing *Listeria* contamination prevalence and concentration on contaminated agents on Wednesday at Midday for Facility A and B baseline conditions by Zone group.** Prevalence of *Listeria* contamination within each area of Facility A (panel A) and Facility B (panel B). Both facilities show higher prevalence in Zone 3 than Zones 1 and 2. Log_10_ concentrations (CFU/cm^2^) of *Listeria* on contaminated agents within each Zone group of Facility A (panel C) and Facility B (panel D) with median concentrations listed showing low level of contamination.

#### Scenario Analysis

**Table S11. Comparisons of median corrective action efficacy against baseline conditions and corresponding interquartile range (IRQ)**

| Scenario model-notation | Median Efficacy (%) (25^th^ percentile-75^th^ percentile) | | | |
| --- | --- | --- | --- | --- |
|  | A | | B | |
|  | Wet | Dry | Wet | Dry |
| PR_01 | 0.0 (-124.9-57.6) | 0.0 (-71.4-46.7) | 0.0 (-16.7-13.6) | 0.0 (-27.6-30.0) |
| PR_02 | 0.0 (-141.0-53.6) | 0.0 (-80.0-45.1) | 0.0 (-18.2-16.7) | 0.0 (-36.3-33.3) |
| PR_03 | 0.0 (-134.6-58.7) | 0.0 (-75.0-46.7) | 0.0 (-20.0-16.7) | 0.0 (-33.3-35.7) |
| LR_01 | 0.0 (-149.9-64.9) | 0.0 (-70.3-47.4) | 0.0 (-9.2-11.3) | 0.0 (-12.1-25.0) |
| LR_02 | 0.0 (-99.5-65.6) | 7.7 (-50.0-50.0) | 0.0 (-6.5-14.3) | 0.0 (0.0-33.3) |
| LR_03 | 0.0 (-99.8-66.0) | 6.7 (-54.0-55.5) | 0.0 (-6.2-14.3) | 0.0 (0.0-40.0) |
| PZ_01 | 0.0 (-122.9-60.0) | 9.5 (-64.4-50.6) | 0.0 (-24.3-18.2) | 0.0 (-33.3-33.3) |
| PZ_02 | 0.0 (-115.7-58.9) | 16.7 (-57.4-57.1) | 0.0 (-20.0-23.5) | 8.7 (-28.9-41.0) |
| PZ_03 | 0.0 (-99.9-62.5) | 28.6 (-28.6-69.2) | 2.9 (-19.4-26.3) | 16.7 (-12.9-50.5) |
| PZ_04 | 0.0 (-100.0-63.6) | 44.4 (0.0-87.5) | 6.7 (-19.4-36.3) | 32.3 (0.0-75.0) |
| LZ_01 | 0.0 (-222.0-45.1) | 18.2 (-50.0-55.5) | 0.0 (-16.7-25.0) | 16.7 (-20.0-50.0) |
| LZ_02 | 0.0 (-122.8-52.9) | 28.6 (-25.0-70.3) | 5.0 (-14.3-25.0) | 28.6 (0.0-66.6) |
| LZ_03 | 0.0 (-145.4-53.6) | 36.3 (-12.5-76.9) | 6.7 (-12.5-33.3) | 33.3 (0.0-72.7) |
| EC_01 | 2.9 (-131.3-64.7) | 0.0 (-66.6-49.9) | 0.0 (-15.8-22.8) | 0.0 (-33.3-32.0) |
| EC_02 | 12.5 (-86.1-81.8) | 6.9 (-57.4-53.4) | 8.6 (-9.6-29.6) | 7.1 (-25.0-33.3) |
| EC_03 | 31.0 (-49.9-90.9) | 17.2 (-54.8-55.5) | 20.0 (0.0-47.0) | 14.3 (-27.6-40.0) |
| EC_04 | 96.1 (49.9-100.0) | 40.0 (-25.0-71.4) | 69.2 (36.3-86.9) | 33.3 (-16.7-66.6) |
| MI_01 | 0.0 (-100.0-52.0) | 0.0 (-54.0-33.3) | 0.0 (0.0-0.0) | 0.0 (0.0-0.0) |
| MI_02 | 9.9 (-133.2-59.3) | 8.3 (-62.5-50.0) | 47.1 (25.0-65.5) | 55.8 (20.0-83.3) |
| AI_01 | 29.3 (-54.0-84.3) | 10.0 (-50.0-55.7) | 25.0 (0.0-49.9) | 0.0 (-20.0-25.0) |
| AI_02C1 | 18.2 (-149.7-58.2) | -30.0 (-149.7-58.2) | N/A | N/A |
| AI_02C2 | 52.8 (-33.3-86.6) | -9.3 (-119.9-42.8) | N/A | N/A |
| AI_02 | N/A | N/A | 33.3 (8.8-50.0) | 49.9 (10.0-75.0) |
| AI_03C1 | 0.0 (-149.9-55.5) | -45.8 (-149.8-14.3) | N/A | N/A |
| AI_03C2 | 0.0 (-203.0-45.5) | -33.3 (-137.4-22.2) | N/A | N/A |
| AI_03 | N/A | N/A | 17.5 (-5.1-40.0) | 0.0 (-34.4-36.3) |
| AI_04 | 45.3 (-83.6-77.7) | 42.8 (-18.2-75.0) | 0.0 (-100.0-50.0) | 84.6 (28.6-99.9) |
| CI_01 | 20.8 (-80.8-81.9) | 36.8 (-14.3-74.9) | 25.0 (0.0-50.0) | 10.0 (-33.3-41.6) |
| CI_02 | 50.0 (-49.9-87.0) | 42.8 (-22.2-73.4) | 4.8 (-76.0-55.0) | 83.3 (33.3-99.9) |
| CI_03 | 45.3 (-88.1-77.8) | 75.0 (37.2-91.6) | 15.4 (-75.3-61.5) | 83.3 (28.6-99.9) |

#### Random Probability Occurrence Reduction

Facility A did not project any changes to median prevalence in either area against the baseline beyond 1.88 percentage point (pp) decrease (Fig S3.A) following an increase in time to random contamination introduction by 25-75%. Median concentrations on contaminated agents showed negligible changes in both the wet and dry area (Fig S3.C).

Facility B also showed no change in wet or dry area median prevalence against the baseline model (Fig S3.B). Changes in median concentration (Fig S3.D) in the respective areas were negligible.

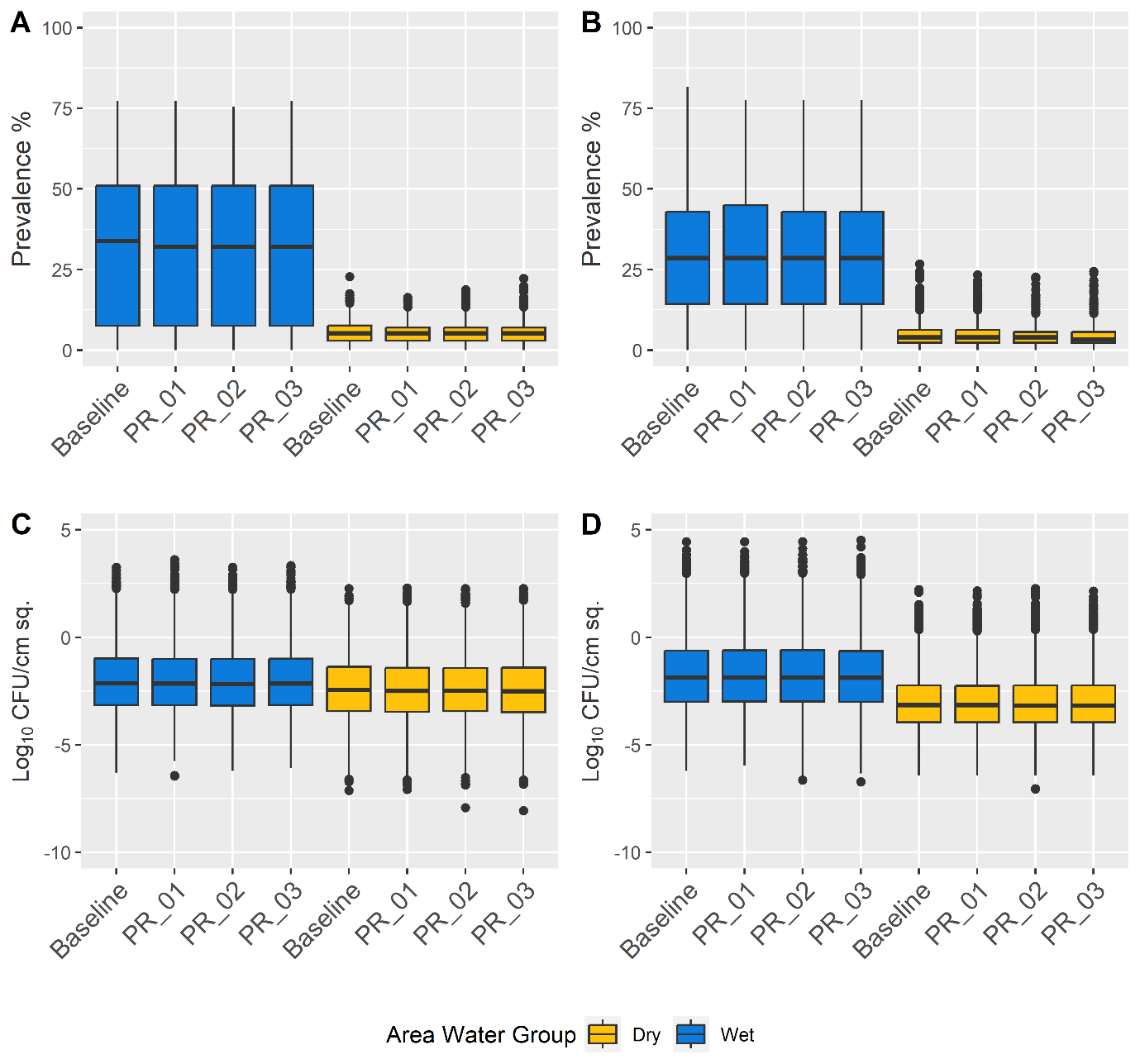

**Fig S3. Boxplots describing the effects of reducing the rate of Random Event Occurrence by 25-75% from baseline on both wet (blue) and dry (yellow) area *Listeria* contamination prevalence and concentration in both models.** Prevalence of *Listeria* contamination of all agents within each area of Facility A (panel A) and Facility B (panel B). Log_10_ concentrations (CFU/cm^2^) of *Listeria* on contaminated agents within each area of Facility A (panel C) and Facility B (panel D).

#### Random Load Reduction

Facility A showed a small decrease in median prevalence across iterations of a simulation in both wet and dry areas against the baseline model (Fig S4.A): the wet area showed a maximum drop of 5.66 pp, while dry areas experienced a maximum drop of 0.58 pp. Median *Listeria* concentration (Fig S4.C) in wet areas showed almost no difference between scenarios (maximum drop of 0.04 log_10_ CFU/cm^2^) and a similarly small decrease in dry areas of 0.07 log_10_ CFU/cm^2^.

Facility B's (Fig S4.B) wet area median prevalence showed a decrease of 4.08 pp at the highest corrective action and dry area median prevalence dropped by 0.57 pp. Median concentration of *Listeria* on positive agents (Fig S4.D) in the wet area decreased by 0.02 log_10_ CFU/cm^2^ and in the dry area by 0.24 log_10_ CFU/cm^2^.

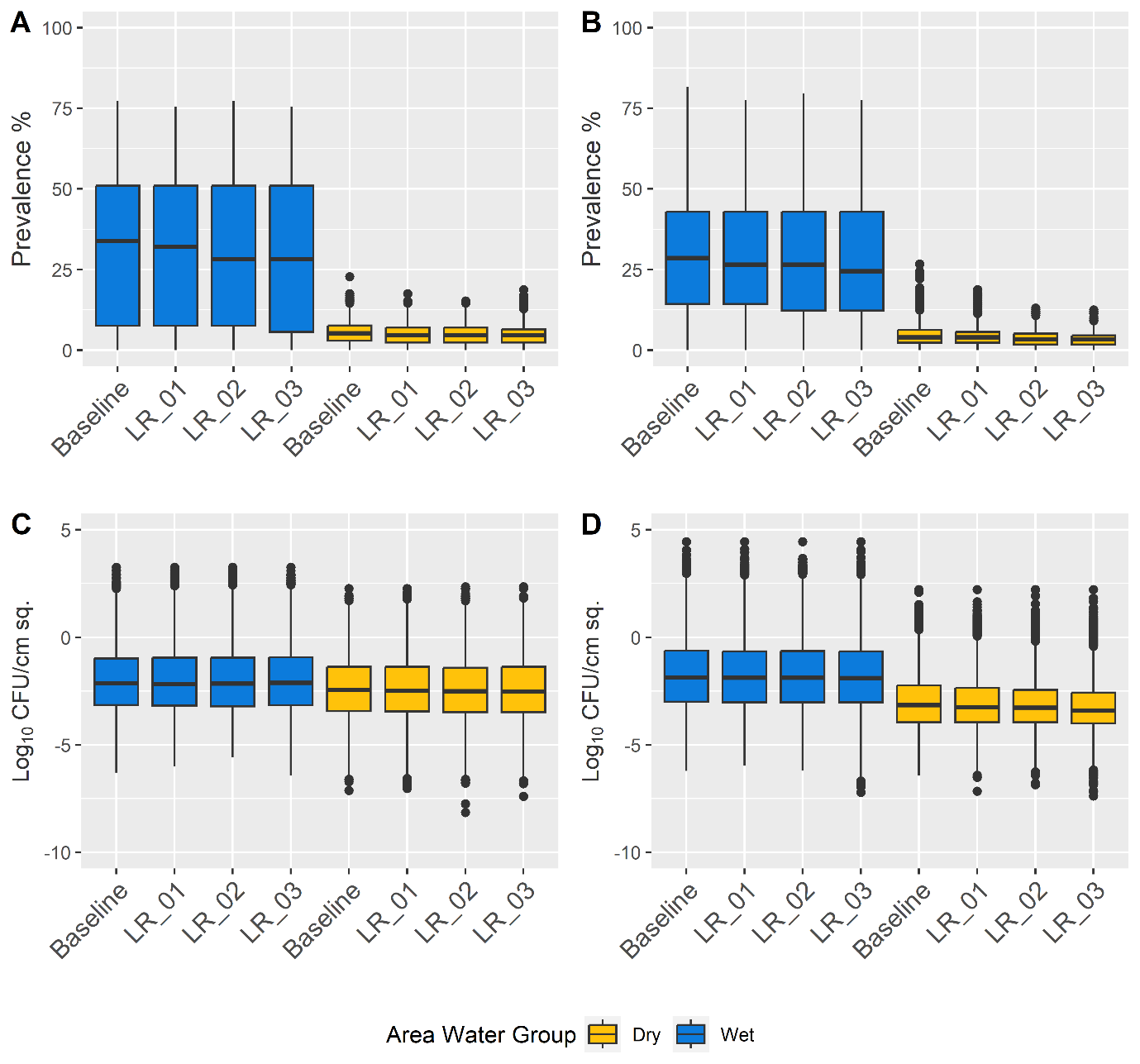

**Fig S4. Boxplots describing the effects of Random Load reduction by 1-3 log_10_ on both wet and dry area contamination prevalence and concentration in both models.** Prevalence of *Listeria* contamination of all agents within each area of Facility A (panel A) and Facility B (panel B). Log_10_ concentrations (CFU/cm^2^) of *Listeria* on contaminated agents within each area of Facility A (panel C) and Facility B (panel D).

#### Z4 Probability Event Reduction

In Facility A, median prevalence in wet and dry areas showed at most a decrease by 2.34 pp at 0% probability of Z4 (Fig S5.A). Median concentrations on positive agents in the wet area showed effectively negligible fluctuation (Fig S5.C), while the median of dry area concentrations increased with each corrective action, reaching a maximum increase of 0.66 log_10_ CFU/cm^2^ from baseline median.

Facility B followed a similar pattern of minimal improvement, with median wet area prevalence decreasing by 2.04 pp at 0% Z4 event probability, and dry area prevalence decreasing by 1.14 pp (Fig S5.B). Median concentration on positive agents in wet areas did not change beyond minor fluctuations, while dry area median concentrations showed a small trend of increasing as probability decreased, culminating in a maximum difference of 0.11 log_10_ CFU/cm^2^ from baseline at 0% (Fig S5.D).

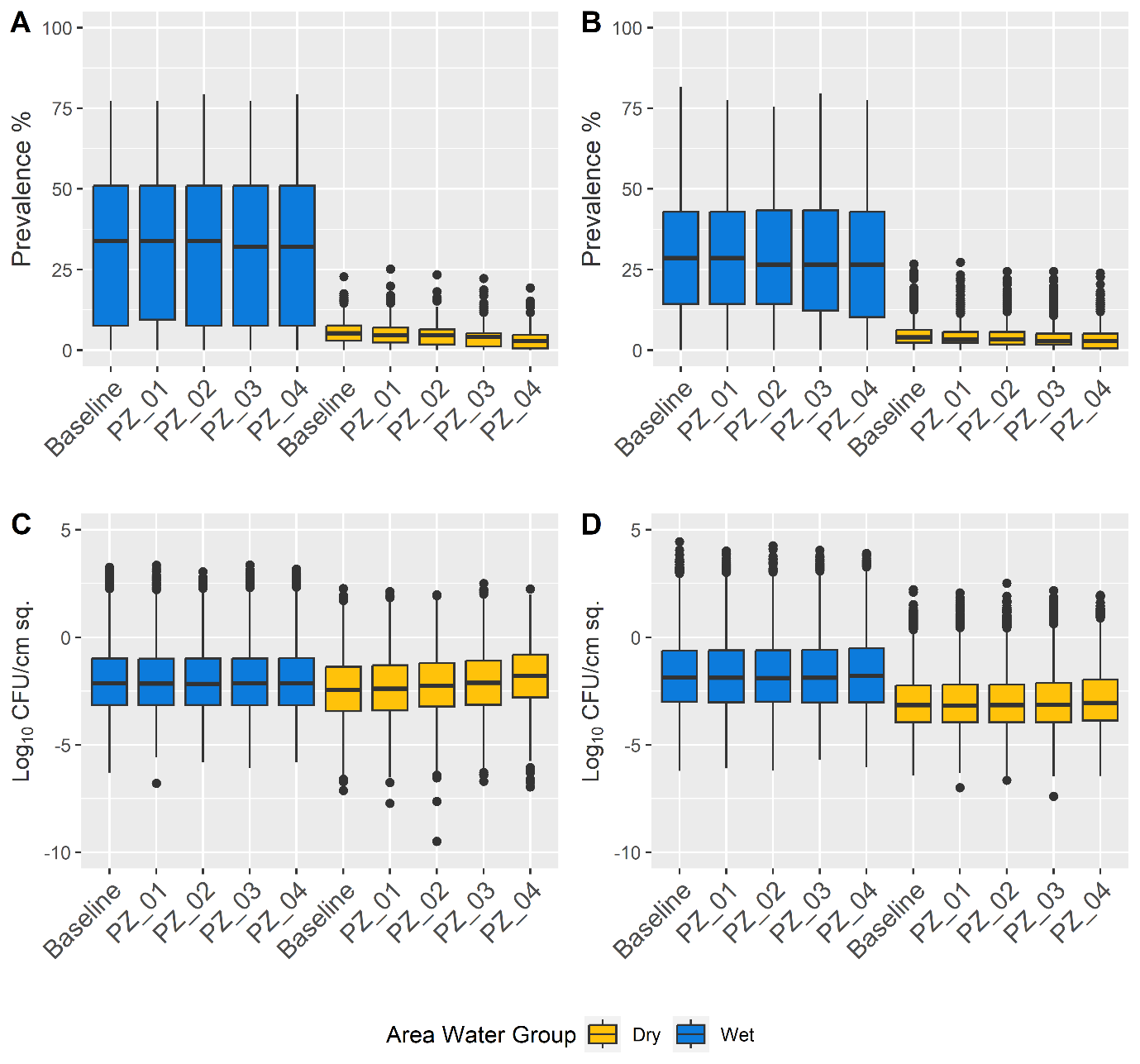

**Fig S5** **Boxplots describing the effects of reducing Z4 Event Occurrence from 100%-0% in increments of 25% on both wet (blue) and dry (yellow) area contamination prevalence and concentration in both models.** Prevalence of *Listeria* contamination of all agents within each area of Facility A (panel A) and Facility B (panel B 1). Log_10_ concentrations (CFU/cm^2^) of *Listeria* on contaminated agents within each area of Facility A (panel C) and Facility B (panel D).

#### Z4 Load Reduction

Facility A showed little-to-no-changes in area prevalence values against the baseline (Fig S6.A) beyond minor fluctuations and a minor decrease in dry area median, dropping by a maximum 0.63 pp at 3 log_10_ Reduction. The median concentration of the wet area showed no major change against the baseline model (0.04 log_10_ CFU/cm^2^ decrease) while dry showed a slight increase in median concentration (0.36 log_10_ CFU/cm^2^) at 3 log_10_ Reduction (Fig S6.C).

Similarly, Facility B's largest log_10_ reduction produced no change to median prevalence in the wet area and a reduction of 1.14 pp in the dry area (Fig S6.B). Median concentration on positive agents in the wet area did not change beyond minor quantities and dry by a maximum of 0.05 log_10_ CFU/cm^2^ (Fig S6.D).

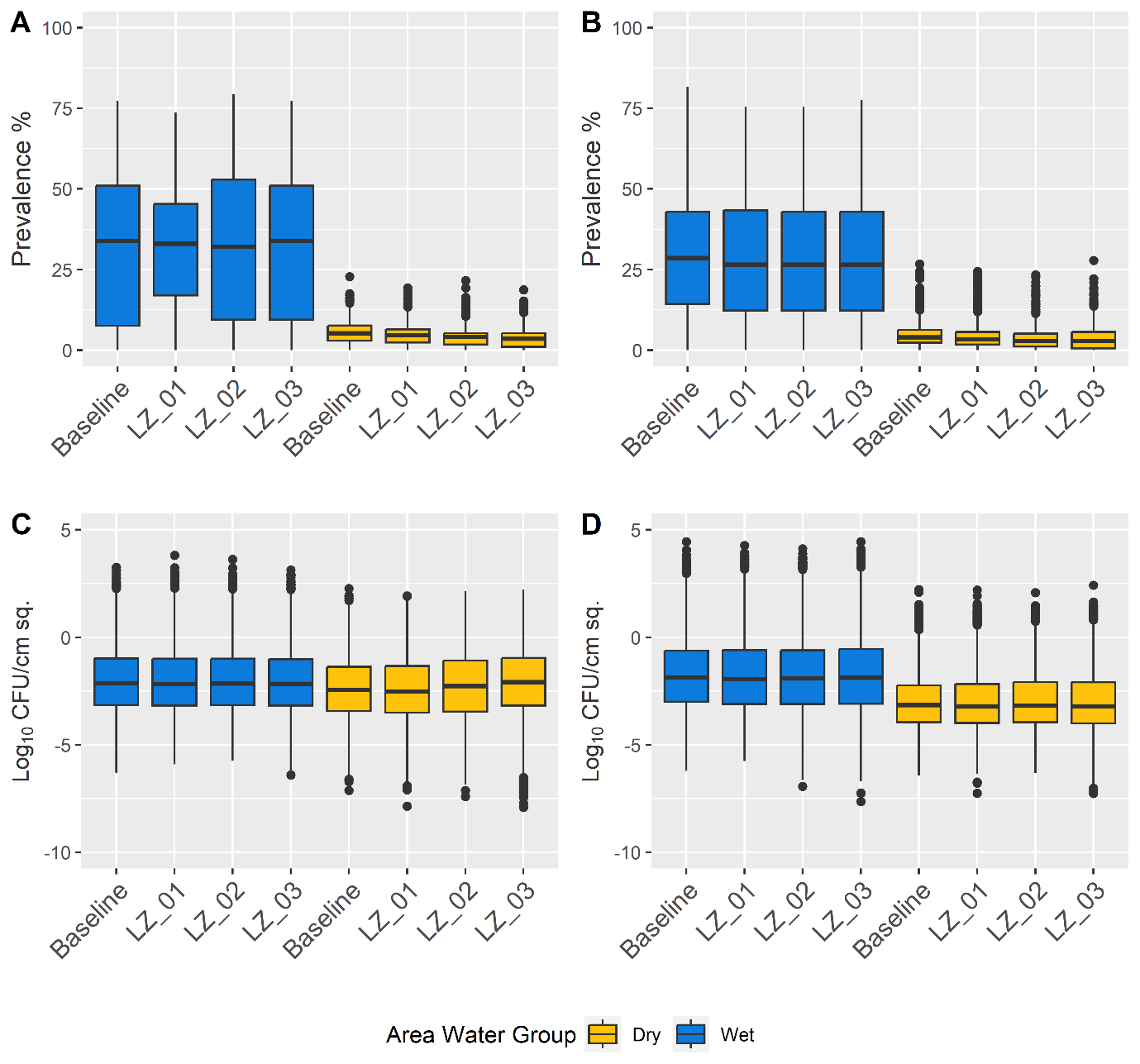

**Fig S6. Boxplots describing the effects of Z4 Load reduction by 1-3 log_10_ on both wet and dry area contamination prevalence and concentration in both models.** Prevalence of *Listeria* contamination of all agents within each area of Facility A (panel A) and Facility B (panel B). Log_10_ concentrations (CFU/cm^2^) of *Listeria* on contaminated agents within each area of Facility A (panel C) and Facility B (panel D).

#### Listeria Prevalence in Incoming Raw Produce Reduction

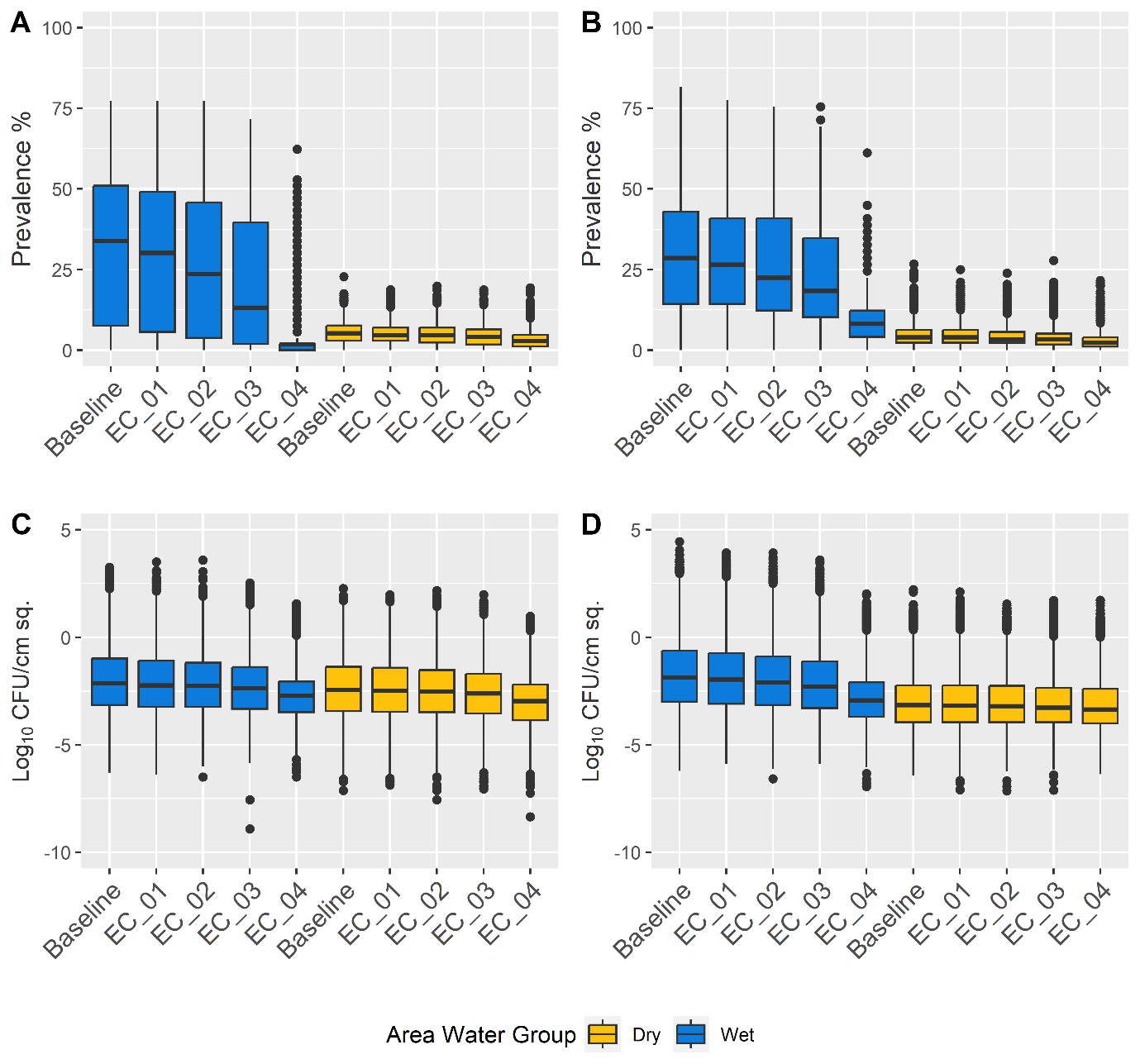

**Fig S7. Boxplots describing the effects of reducing prevalence in incoming produce on both wet (blue) and dry (yellow) area agents.** *Listeria* prevalence in incoming produce was reduced by multiplying the baseline *Listeria* prevalence by the factors of 0.75, 0.5, 0.25 and 0 (scenarios EC_01, EC_02, EC_03, and EC_04, respectively); this simulates produce being treated prior to arriving in the packinghouse packing room. A: Facility A *Listeria* contamination prevalence of all agents in wet and dry areas. B: Facility B *Listeria* contamination prevalence of all agents in wet and dry areas. C: Facility A *Listeria* log_10_ concentrations on all positive agents in wet and dry areas. D: Facility B *Listeria* log_10_ concentrations on all positive agents in wet and dry areas.

#### Cleaning Effectiveness Improvement and Weekend Deep Clean

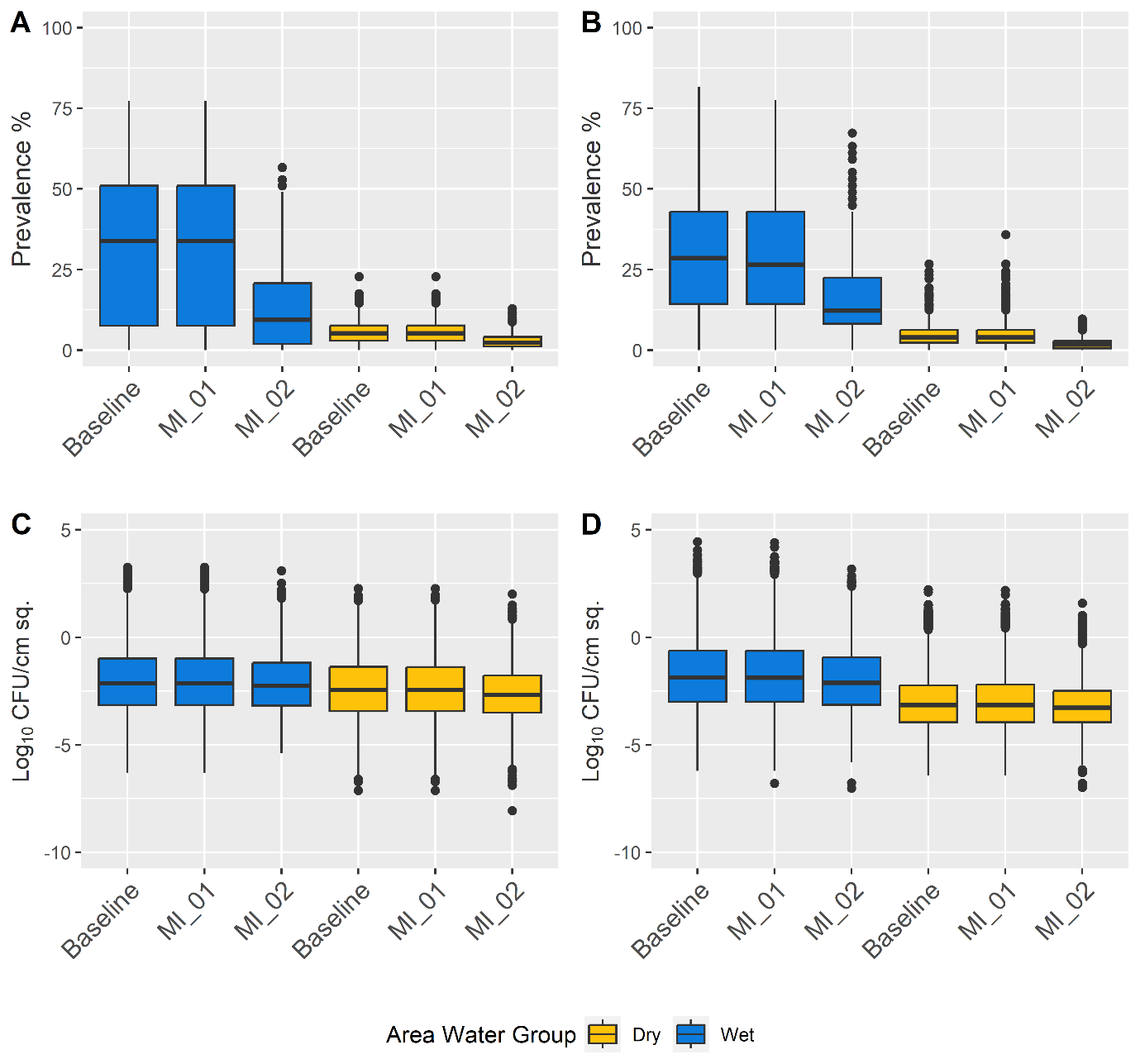

**Fig S8.** **Boxplots describing the effects of increasing *Listeria* removal by 3 log_10_ (scenario MI_01) or performing weekend deep cleaning (scenario M1_02) on both wet (blue) and dry (yellow) area agents.** Prevalence of *Listeria* contamination of all agents within each area of Facility A (panel A) and Facility B (panel B). Log_10_ concentrations (CFU/cm^2^) of *Listeria* on contaminated agents within each area of Facility A (panel C) and Facility B (panel D). (MI_01: Cleaning Effectiveness Improvement (*Listeria* removal during reduction events increased by 3 log_10_); MI_02: Weekend Deep Clean (Removal of all *Listeria* from all agents regardless of cleanability status every Sunday))

#### Enhanced Flume Water Treatment, Broad and Directed Model-based Master Sanitation Schedule Restructuring, and Transmission Pathways Modification Corrective Action

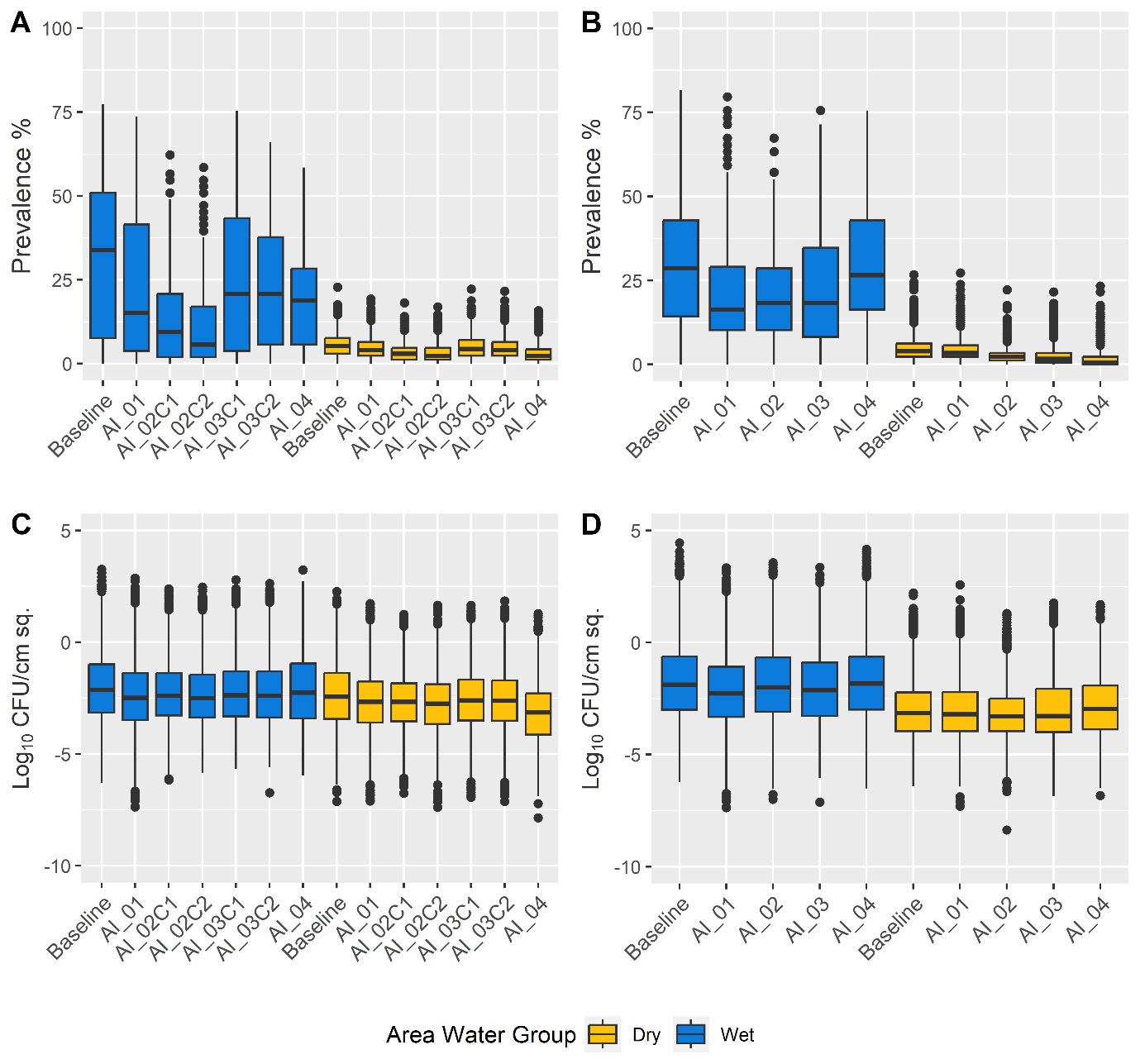

**Fig 9.** **Boxplots describing the effects of various agent-specific corrective actions (scenarios AI_01, AI_02/AI_02C1/AI_02C2, AI_03/AI_03C1/AI_03C2, AI_04) on both wet (blue) and dry (yellow) area agents.** Prevalence of *Listeria* contamination of all agents within each area of Facility A (panel A) and Facility B (panel B). Log_10_ concentrations (CFU/cm^2^) of *Listeria* on contaminated agents within each area of Facility A (panel C) and Facility B (panel D). (AI_01: Enhanced Flume Water Treatment (2 log_10_ removal of *Listeria* in flume agent per hour of production); AI_02/AI_02C1/AI_02C2: Broad Model-based Master Sanitization Schedule Restructuring (Agent cleaning and sanitation schedules were fully reassigned according to mean contamination probability; Facility A was given a daily schedule for both “Cleaning Only” and “Cleaning & Sanitation” respectively); AI_03/AI_03C1/AI_03C2: Directed Model-based Master Sanitization Schedule Restructuring (Sanitization of agents with a mean predicted contamination probability ≥66% in the baseline model was set to daily cleaning; Facility A was given a daily schedule for both “Cleaning Only” and “Cleaning & Sanitation” respectively); AI_04: Transmission Pathways Modification Corrective Action (Facility A: Drain compartmentalization; Facility B: Separation of forklift area assignment))

#### Combined Corrective Actions 01, 02, and 03

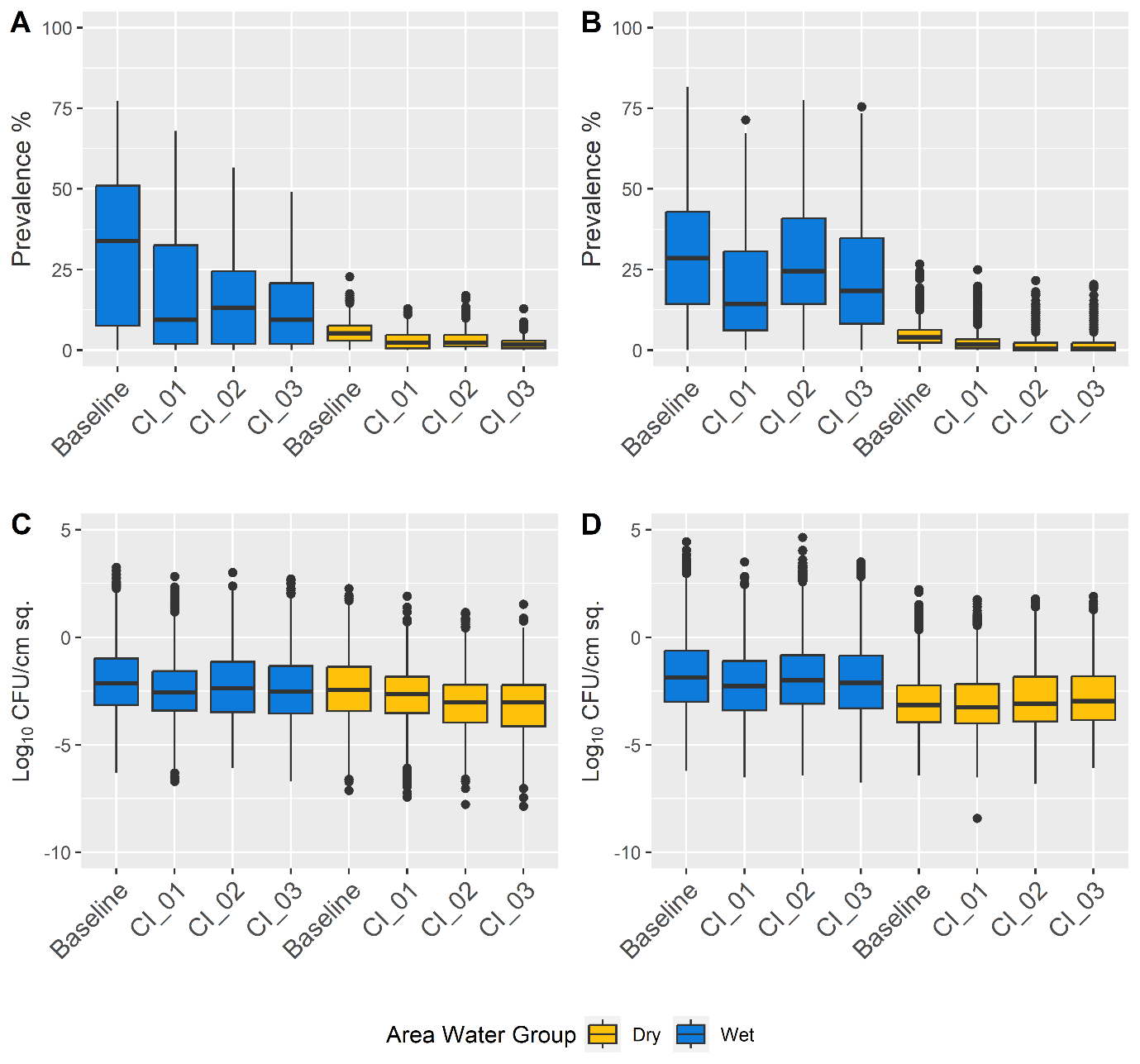

**Fig S10.** **Boxplots describing the effects of the combined synergistic effect of simultaneous actions (scenarios CI_01, CI_02, CI_03) on both wet (blue) and dry (yellow) area agents.** Prevalence of *Listeria* contamination of all agents within each area of Facility A (panel A) and Facility B (panel B). Log_10_ concentrations (CFU/cm^2^) of *Listeria* on contaminated agents within each area of Facility A (panel C) and Facility B (panel D). (CI_01: 50% Reduction of *Listeria* Prevalence in Incoming Produce and Broad Model-based Master Sanitization Schedule Restructuring were applied simultaneously in Facility A; 50% Reduction of *Listeria* Prevalence in Incoming Produce and Directed Model-based Master Sanitization Schedule Restructuring were applied simultaneously to Facility B; CI_02: 50% Reduction of *Listeria* Prevalence in Incoming Produce and Transmission Pathways Modification Corrective Action (Facility A: Drain compartmentalization; Facility B: Separation of forklift area assignment) were applied simultaneously to both; CI_03: Broad Model-based Master Sanitization Schedule Restructuring and Transmission Pathways Modification Corrective Action (Facility A: Drain compartmentalization; Facility B: Separation of forklift area assignment) were applied simultaneously in Facility A; Directed Model-based Master Sanitization Schedule Restructuring and Transmission Pathways Modification Corrective Action (Facility A: Drain compartmentalization; Facility B: Separation of forklift area assignment) were applied simultaneously to Facility B)
